# *Schistosoma mansoni* infections are associated with hepatosplenic organometry within the context of repeated praziquantel treatment and co-endemic infections of malaria, hepatitis B, and HIV: a population-based study in rural Uganda

**DOI:** 10.64898/2026.03.06.26347797

**Authors:** Kayla Seggelke, Max M. Lang, Betty Nabatte, Victor Anguajibi, Benjamin Ntegeka, Tymothy Mugume, Simon Mpooya, Narcis B. Kabatereine, Adrian D. Smith, Goylette F. Chami

## Abstract

**Background:** *Schistosoma mansoni* is a leading cause of hepatosplenic disease in sub-Saharan Africa. Yet, associations with current *Schistosoma mansoni* infections and hepatosplenic organometry remain unclear in the context of widespread mass drug administration and co-endemic infections.

**Methods:** From January to February 2024, we conducted a community-based, cross-sectional study nested within the SchistoTrack cohort in three districts of Uganda. Liver and spleen dimensions were assessed via point-of-care B-mode ultrasound for 3121 individuals. Organ dimensions were classified using the standard deviations from height-standardized internal reference values derived from an infection-free population. Multinomial logistic regressions were run for children (5–17 years) and adults (18+ years) separately. Population attributable fractions (PAFs) were used to estimate the proportion of abnormal organometry statistically attributable to each infection. Key exposures were *S. mansoni*, malaria, human immunodeficiency virus (HIV), and hepatitis B virus (HBV) alongside a comprehensive set of social, biomedical, and other covariates controlled for.

**Results:** Moderate-to-severe splenic enlargement was observed in 29.1% (438/1507) of children and 23.3% (376/1614) of adults. Among adults, 20.9% (337/1614) had left liver lobe enlargement and 18.8% (303/1614) had right liver lobe shrinkage. In children, severe splenic enlargement was statistically attributable to malaria (PAF 46.7%; Relative Risk Ratio (RRR) 3.96, 95% CI 2.64–5.92) and *S. mansoni* infection intensity (PAF 23.6%; RRR 1.12, 95% CI 1.04–1.20). In adults, *S. mansoni* intensity was associated with moderate left liver lobe enlargement (PAF 12.4%; RRR 1.11, 95% CI 1.04–1.18). In adults, HIV was associated with severe left liver lobe shrinkage (RRR 4.50, 95% CI 1.19–17.00) and severe splenomegaly (RRR 3.62, 95% CI 1.58–8.33), while HBV was associated only with severe left liver lobe shrinkage (RRR 2.54, 95% CI 1.07–6.03). Praziquantel treatment in the past year showed inconsistent associations and no clear protective pattern.

**Conclusion:** Current *S. mansoni* infection intensity remains associated with splenomegaly in children despite controlling for concurrent malaria positivity, and with hepatomegaly in adults despite HIV and HBV associations.

## Introduction

Schistosomiasis is endemic in 41 countries in sub-Saharan Africa (SSA), where an estimated 237 million people require treatment for current schistosome infections [1]. Hepatosplenic schistosomiasis caused by *Schistosoma mansoni* is common in SSA. For example, the prevalence of hepatosplenic schistosomiasis has been estimated to be as high as 55% in children and adults in rural communities in Uganda [2]. The eggs produced by the female parasitic flukes induce granulomatous inflammatory responses that, if uncontrolled by the host, may lead to periportal fibrosis (PPF), including portal vasculature restructuring. Changes in spleen and liver sizes often precede or accompany PPF [2, 3]. It remains unknown whether current *S. mansoni* infection correlates with hepatosplenic organometry, given the widespread availability of praziquantel through mass drug administration (MDA) and co-endemic infections of malaria, human immunodeficiency virus (HIV), and hepatitis B (HBV).

The assessment of organometry, mostly focused on hepatosplenomegaly, has been used for early indications of disease in children due to the heaviest burden of infection being concentrated in this group [4–7]. However, recently, it has been shown that there is a disconnect between more severe hepatosplenic outcomes, in particular PPF that accompanies easily observed changes in organometry, and current intestinal schistosome infection [3]. The highest burden of PPF has been shown to be in adults, supporting the need for additional studies within this age group that also rely on community-based sampling rather than school-based sampling [3]. It is assumed that schistosomiasis rarely causes cirrhosis or cirrhosis-like outcomes, including shrunken livers [8], yet few studies have examined these severe outcomes.

There is no agreement as to the direction and significance of organometry and current schistosome infections within the context of routine praziquantel treatment or MDA. For instance, while some studies report significant associations between *S. mansoni* and hepatosplenomegaly [4, 7], others have found no significant difference in hepatosplenomegaly based on infection status [4, 5, 9, 10]. Detectable morbidity may depend on infection intensity, as several studies note that hepatosplenomegaly was only significantly associated with *S. mansoni* in individuals with heavy infections [11–13]. Yet, for related outcomes of PPF, there is no association of current intestinal schistosome infections and PPF within the context of routine MDA [14]. The lack of morbidity management guidelines for schistosomiasis, coupled with the inability of MDA to resolve schistosomiasis-associated morbidities, highlights the need to revisit the use of routine organometry for assessing schistosomiasis-related morbidity in endemic countries.

Schistosomiasis often is co-endemic with malaria, HIV, and HBV in rural SSA [15–18]. Malaria and schistosome co-infections are most often studied with respect to splenomegaly only [7, 19, 20]. Yet, there is a disagreement as to whether splenomegaly should be assessed for schistosomiasis in areas endemic with malaria. The World Health Organization (WHO) protocol for assessing schistosomiasis morbidity (Niamey Protocol) recommends against assessing splenomegaly in malaria endemic areas with the assumption that malaria is the sole cause [21]. Yet, it has been shown that malaria infection facilitates schistosome-initiated splenomegaly [6].

HIV and HBV have been shown to independently correlate with outcomes related to cirrhosis, and as such, shrunken livers [22, 23], whether schistosome infections independently contribute to this outcome or exacerbate HIV and HBV pathology remains to be explored. To our knowledge, no studies examine both schistosome and either HIV or HBV infections and associated organometry. A recent meta-analysis found that current schistosome infection is not associated with PPF in the context of MDA, suggesting that current infection status may not be the best proxy for cumulative morbidity or severe fibrosis/cirrhosis risk [14], though HIV infection has been shown to be linked to PPF [2, 3].

Concerning the effect of praziquantel on organometry, the evidence is inconclusive. Vennervald et al. [24] demonstrated regression of early hepatosplenomegaly in children following praziquantel treatment. Yet, in the context of routine MDA, the impact of one round of praziquantel on the likelihood of improved liver or spleen organometry is negligible with small effect sizes [25].

We conducted a study of organometry using point-of-care B-mode ultrasound on 3121 individuals aged 5–90 years in the SchistoTrack cohort in rural Uganda. Reference tables for abnormal organometry were constructed. We identified risk factors for splenic enlargement, left liver lobe shrunkenness/enlargement, and right liver lobe shrunkenness/enlargement. Detailed covariates on sociodemographics, water, sanitation, and hygiene (WASH) conditions, treatment history, and behavioral risk factors were measured in addition to the key exposures of *S. mansoni*, malaria, HIV, and HBV. The aim of this study was to assess the associations of current schistosome infections with abnormal organometry in the context of widespread MDA and co-endemic infections.

## Methods

### Study design and participant sampling

This cross-sectional study was nested within the SchistoTrack cohort during the 2024 timepoint. SchistoTrack is a community-based prospective cohort established in 2022 across three rural districts in Uganda: Pakwach (along the Albert Nile), Buliisa (Lake Albert), and Mayuge (Lake Victoria) [26]. Households were selected by random sampling within villages. One adult (≥18 years) and one child (5–17 years) were selected by the household head for clinical assessment. Cohort details and sampling are further described by Puthur et al. [26].

### MDA history in study area

In these districts, annual school-based and community-based MDA with praziquantel was started in 2003 to target eligible individuals aged at least five years, except in 2019 (all districts) and 2022–2023 (Mayuge), when only school-based MDA was completed [3, 27]. As reported to the WHO Expanded Special Project for the Elimination of Neglected Tropical Diseases [27], of the 21 possible annual rounds of MDA before this study, Buliisa and Pakwach districts received only 14 rounds, whereas Mayuge district received 17 rounds. The most recent round of MDA in our study districts was in 2023 (Mayuge) and 2022 (Buliisa and Pakwach). The mean administrative treatment coverage for community-based MDA was 80 · 8% in Buliisa, 80 · 6% in Pakwach, and 76 · 2% in Mayuge. Among all rounds of MDA implementation, Pakwach and Buliisa districts missed WHO-suggested targets of 75% treatment coverage by more than 10% for only 1 year, whereas Mayuge district missed this target by more than 10% for 3 non-consecutive years. All study districts had no MDA in 2007 and 2008.

### Organometry outcomes

Abdominal ultrasound exams were performed to assess organometry. Philips Lumify C5-2 curved linear array transducers with Lenovo 8505-F tablets with Android 9 Pie were used. Sonographers were blinded to all infection and clinical outcomes not assessed via ultrasound. The left liver lobe was measured via a longitudinal section at the parasternal line, using the abdominal aorta as reference. The right liver lobe was measured from the midclavicular line, using the gall bladder as reference. Spleen lengths for normal rounded shapes were measured between the two poles, crossing through the splenic hilum; for distorted spleen shapes, the length was measured past the splenic hilum to avoid underestimation due to organ curvature [3].

To define abnormalities and account for anthropometric variation, height-standardized reference values were constructed. Among all study participants, a healthy reference population was identified. Participants had to be negative for *S. mansoni*, malaria, HBV, and HIV; not pregnant; to have reported no current (within the past 12 months) alcohol consumption; and to have no liver fibrosis as diagnosed via ultrasound. Height deciles were constructed for this healthy population. Abnormal organometry for all study participants was identified in comparison to the number of standard deviations (SDs) of organ size from the mean of organ size within the same height decile of the healthy reference population. Liver outcomes included five categories: severely shrunken (≤ −2 SD), moderately shrunken (> −2 to ≤ −1 SD), normal (−1 to +1 SD), moderately enlarged (≥ +1 to < +2 SD), and severely enlarged (≥ +2 SD). Splenic outcomes included three categories: normal, moderately enlarged (≥ +1 to < +2 SD), and severely enlarged (≥ +2 SD).

### Exposures

Key exposures included *S. mansoni*, malaria, HBV, and HIV. For *S. mansoni*, two slides from one stool sample were prepared for Kato–Katz microscopy as described elsewhere [3]. Infection intensity was defined as natural log-transformed eggs per gram (EPG) plus one. Finger-prick blood samples were collected for rapid diagnostic testing (RDT) to assess malaria (Bioline Malaria Ag P.f/Pan RDT, Abbott), HBV (HBsAg RDT, Abbott), and HIV (Determine HIV-1/2 RDT, Abbott) status. Nurses asked study participants whether they had received praziquantel within the past year from the village health teams outside of the study. Detailed covariate definitions for sociodemographics, WASH, and behavioral risk factors are described in Supplementary Table S6 and were selected based on variables identified as relevant for hepatosplenic schistosomiasis, in particular PPF [3].

### Statistical analysis

All statistical analyses were performed using R v4.2.1. Multinomial logistic regression was used to estimate Relative Risk Ratios (RRR) and 95% confidence intervals (CIs) for all outcomes with one regression each for the spleen, left liver lobe, and right liver lobe. Children and adults were analyzed in separate models to account for physiological differences in organ growth and risk profiles. Base models with key exposures *S. mansoni*, malaria, HIV, and HBV, as well as age, sex, and district, were constructed with HIV and HBV only in models for adults due to the limited prevalence (<5%) in children. Appropriateness of a multinomial model compared to a proportional-odds model was assessed with the Brant test [28]. For variable selection, variables with less than 5% frequency in the study population were excluded. Candidate covariates were added individually to base models and tested using likelihood ratio tests (LRT); variables providing a significant improvement in model fit (*p* < 0.05) were retained. To facilitate direct comparison of risk factors across different organs, the final multivariable models for each population (children and adults) consisted of the union of all significant variables identified for any of the three organ outcomes, respectively. Collinearity was assessed using generalized variance inflation factors (GVIF) implemented in the car package [29, 30].

Sensitivity analyses were performed using multinomial regression on collapsed outcome categories (e.g. any enlargement, normal, or any shrunkenness), and agreement between left and right liver lobe size classifications was assessed using Chi-squared tests. Associations between individual infections and any hepatosplenomegaly outcome were assessed using logistic regression. Additionally, for children, the spleen enlargement model was re-run by substituting binary malaria RDT status with continuous log-transformed parasite density from microscopy readings as defined in Lang et al. [31].

Population attributable fractions (PAFs) were estimated using the G-computation framework [32] to quantify the disease burden statistically attributable to each key infection. PAFs were estimated only for exposures that showed statistically significant associations (*p* < 0.05) in the multivariable models. The PAF represents the estimated percentage reduction in the prevalence of abnormal organometry that would occur if an infection were eliminated from the population. These values were derived by comparing the observed study population to hypothetical counterfactual scenarios. For each infection, a counterfactual population was constructed by setting the exposure for all participants to the reference level, zero EPG for *S. mansoni*, malaria-negative, HIV-negative, or HBV-negative, while retaining observed values for all other covariates in the adjusted models. The PAF was then calculated as the proportional reduction in the mean predicted probability of the outcome in the counterfactual scenario compared to the observed prevalence. The resulting percentages represent the proportion of cases avoidable in the absence of that specific exposure. Uncertainty for these estimates was assessed via 95% CIs derived from 1000 bootstrap iterations.

To validate the construction of the internal reference population, we compared organometric classifications against the reference standards defined by the WHO Niamey Protocol [21, 33]. Participants were re-classified using the fixed height-dependent cut-offs provided in the Niamey guidelines (Annex C) [21]. We calculated the prevalence of abnormal organometry under both classification systems and assessed discordance to determine if external standards systematically over- or under-estimated specific pathologies within the SchistoTrack cohort.

## Results

### Organometry outcomes and infection prevalence

A total of 3302 out of 3385 clinical participants underwent ultrasound examinations in 2024, with 3121 participants with complete data and the focus of analyses with 1507 children and 1614 adults (Figure 1). Participant characteristics are detailed in Table 1 and Supplementary Table S3. The median age was 39 years (Interquartile range (IQR) 30–50) for adults and nine years (IQR 7–12) for children. The majority of adults were female (1051/1614; 65%), whereas the sex distribution was more balanced among children (670/1507; 44% female). Praziquantel uptake through MDA in the past year was low, reported by 20% (325/1614) of adults and 24% (358/1507) of children.

**Fig 1.**
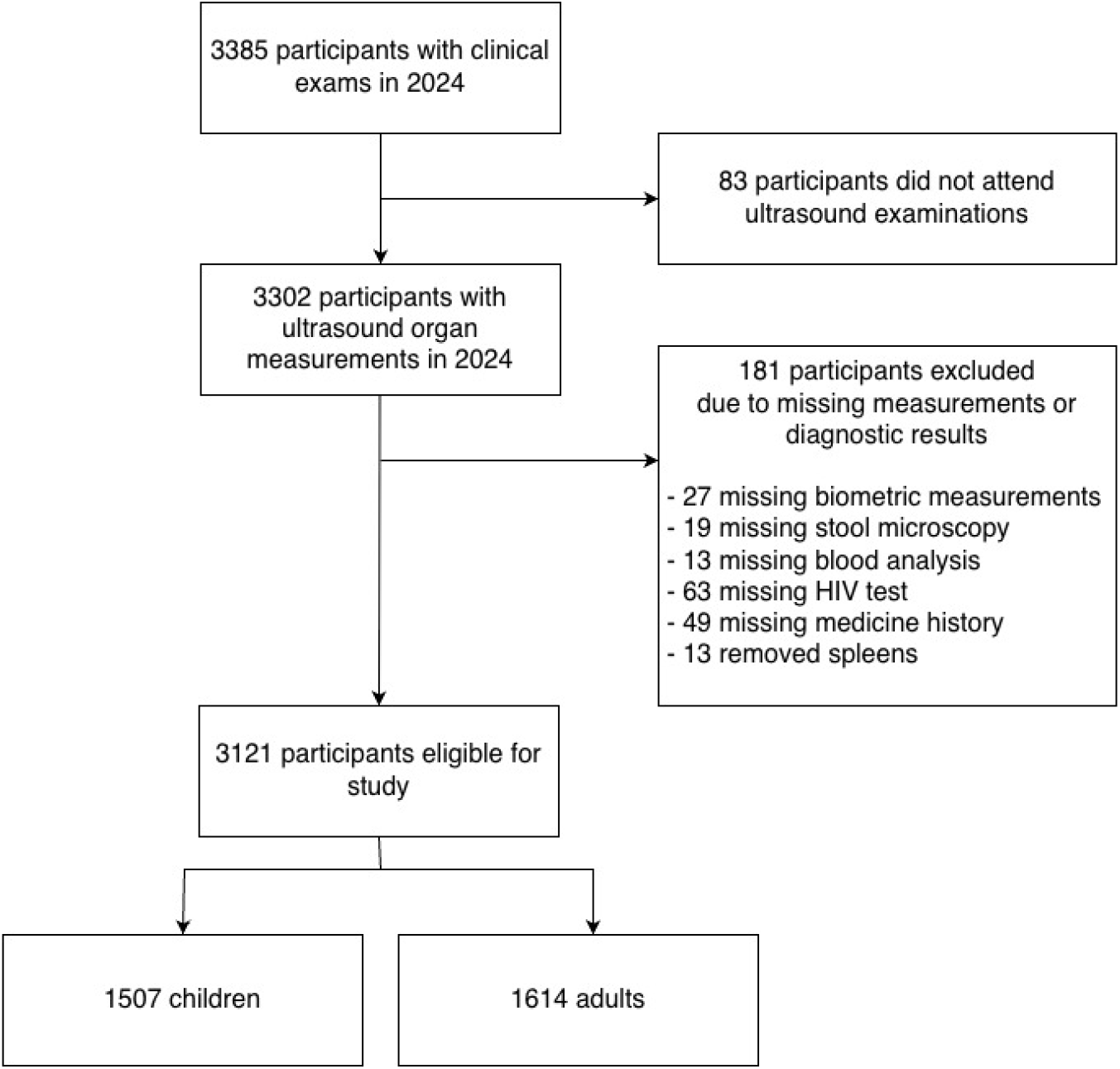
Flowchart of study participant selection. The diagram illustrates the derivation of the analytical sample (*n* = 3121) from the total clinical participants recruited in the SchistoTrack cohort (*n* = 3385) during January–February 2024. Exclusions were made for missing ultrasound examinations or incomplete covariate data. Participants may be missing more than one of the listed sources. The final population was stratified into children (aged 5–17 years; *n* = 1507) and adults (aged ≥18 years; *n* = 1614).

**Table 1.** Demographic, clinical, and parasitological characteristics of the study population. Values are presented as n (%) for categorical variables and median (IQR) for continuous variables.

| Variable | Adults<br>(N = 1614) | Children<br>(N = 1507) |
| --- | --- | --- |
| *S. mansoni* (Positive) | 478 (30%) | 878 (58%) |
| Malaria (Positive) | 211 (13%) | 697 (46%) |
| HBV (Positive) | 140 (8.7%) | 22 (1.5%) |
| HIV (Positive) | 138 (8.6%) | 19 (1.3%) |
| **Left liver lobe** |  |  |
| Severely shrunken | 43 (2.7%) | 27 (1.8%) |
| Moderately shrunken | 156 (9.7%) | 182 (12%) |
| Normal | 1078 (67%) | 1084 (72%) |
| Moderately enlarged | 254 (16%) | 177 (12%) |
| Severely enlarged | 83 (5.1%) | 37 (2.5%) |
| **Right liver lobe** |  |  |
| Severely shrunken | 73 (4.5%) | 63 (4.2%) |
| Moderately shrunken | 230 (14%) | 195 (13%) |
| Normal | 1062 (66%) | 1096 (73%) |
| Moderately enlarged | 209 (13%) | 144 (9.6%) |
| Severely enlarged | 40 (2.5%) | 9 (0.6%) |
| **Spleen** |  |  |
| Normal | 1238 (77%) | 1069 (71%) |
| Moderately enlarged | 216 (13%) | 297 (20%) |
| Severely enlarged | 160 (9.9%) | 141 (9.4%) |
| **Age (years)** | 39.0 (30.0, 50.0) | 9.0 (7.0, 12.0) |
| **Sex (Female)** | 1051 (65%) | 670 (44%) |
| **ART** | 110 (6.9%) | 13 (0.9%) |
| **Praziquantel in past year** | 325 (20%) | 358 (24%) |
| **Antimalarials** | 383 (24%) | 482 (32%) |
| **Alcohol intake** | 322 (20%) | - |
| **Smoking status** | 226 (14%) | - |
| **Years of education** | 4.0 (1.0, 6.0) | 2.0 (1.0, 3.0) |
| **Occupation** |  |  |
| None/other | 884 (55%) | - |
| Farmer | 510 (32%) | - |
| Fisherman | 220 (14%) | - |
| **Water contact** | 1393 (86%) | 1301 (86%) |
| **Tribe** |  |  |
| Alur | 992 (61%) | 944 (63%) |
| Bagungu | 147 (9.1%) | 150 (10.0%) |
| Musoga | 201 (12%) | 160 (11%) |
| Other | 274 (17%) | 253 (17%) |
| **Religion (majority)** | 1212 (75%) | 1154 (77%) |
| **Enrolment year** |  |  |

**Table 1.** Table continued from previous page
|  |  |  |
| --- | --- | --- |
| 2022 | 1035 (64%) | 943 (63%) |
| 2023 | 363 (22%) | 350 (23%) |
| 2024 | 216 (13%) | 214 (14%) |
| **Home quality score** | 3.0 (3.0, 9.0) | 3.0 (3.0, 9.0) |
| **Water purification** | 361 (22%) | 339 (22%) |
| **Hygiene** | 133 (8.2%) | 129 (8.6%) |
| **Sanitation** | 924 (57%) | 866 (57%) |
| **Home ownership** | 1420 (88%) | 1322 (88%) |
| **Years in village** | 18.0 (8.0, 30.0) | 17.0 (8.0, 30.0) |
| **District** |  |  |
| Mayuge | 405 (25%) | 335 (22%) |
| Buliisa | 525 (33%) | 507 (34%) |
| Pakwach | 684 (42%) | 665 (44%) |

Organ dimensions derived from the healthy reference population are presented in Table 2. Abnormality prevalence varied by age group and organ (Table 1). Splenomegaly was common among children: moderate splenic enlargement was observed in 20% (297/1507) and severe enlargement in 9.4% (141/1507) of children. Moderate liver abnormalities affected approximately 10–13% of children, specifically moderate shrinkage (Left lobe 12%, 182/1507; Right lobe 13%, 195/1507) and moderate enlargement (Left lobe 12%, 177/1507; Right lobe 9.6%, 144/1507). Severe liver pathology was less frequent; severe shrunkenness was identified in 1.8% (27/1507) and 4.2% (63/1507) for the left and right lobes, respectively. Severe enlargement was found in 2.5% (37/1507) of left lobes and 0.6% (9/1507) of right lobes.

**Table 2.**
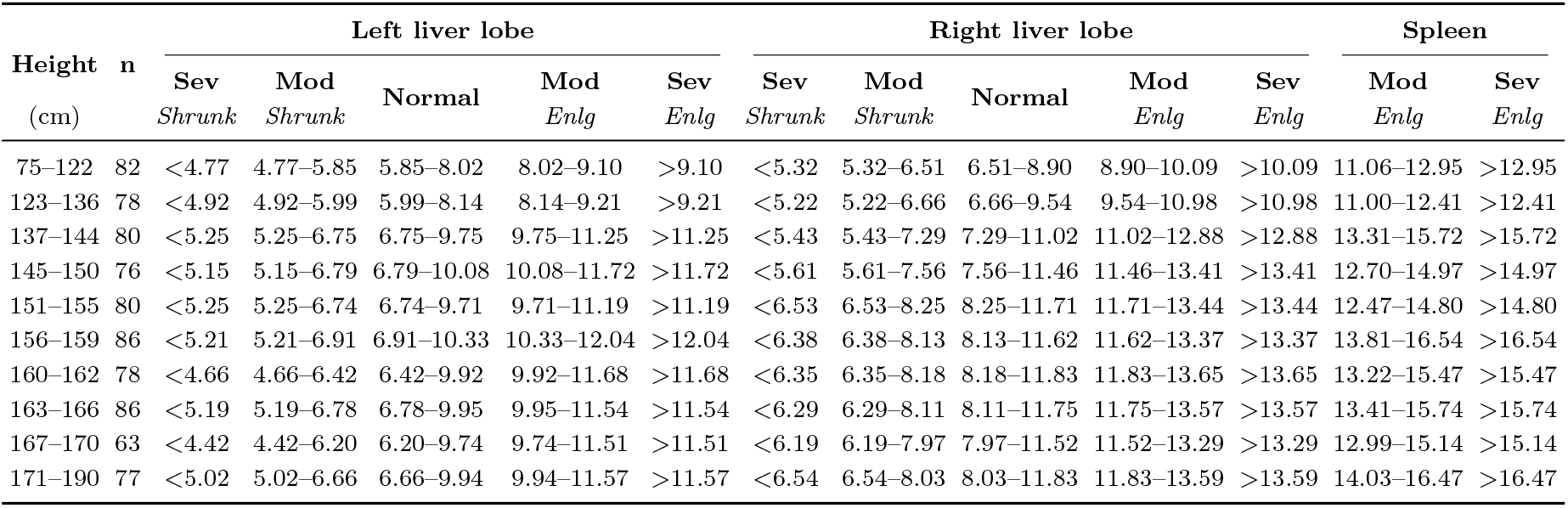
Combined reference values for liver and spleen size classes by height. All measurements are in cm. Values denote the range inclusive of the boundaries. Abbreviations: Sev, Severe abnormality (> 2 SD or < −2 SD); Mod, Moderate abnormality (1 to 2 SD or −1 to −2 SD); Enlg, Enlarged; Shrunk, Shrunken.

Among adults, splenic enlargement was also common, with 13% (216/1614) classified as moderately enlarged and 9.9% (160/1614) as severely enlarged. Moderate liver abnormalities were frequent; 9.7% (156/1614) and 14% (230/1614) had left and right lobe shrunkenness, respectively, while moderate enlargement was observed in 16% (254/1614) for the left lobe and 13% (209/1614) for the right lobe. Severe abnormalities were less common; severe shrunkenness affected 2.7% (43/1614) of left lobes and 4.5% (73/1614) of right lobes, while severe enlargement was identified in 5.1% (83/1614) of left lobes and 2.5% (40/1614) of right lobes. Differences in infection prevalence were observed between children and adults (Table 1 and Table S4). *S. mansoni* prevalence was significantly higher in children (58%, 878/1507) compared to adults (30%, 478/1614; *χ*^2^ = 259.1, *p* < 0.001). Overall infection prevalence also varied by district (Supplementary Table S8), as Pakwach had higher prevalence of S. mansoni and malaria than Mayuge, while HBV and HIV were modestly more common in Buliisa and Pakwach.

### Determinants of abnormal organometry for children

Multinomial models of abnormal organometry with selected covariates are presented in Figures 2, 3, and 4. For splenic morbidity in children, *S. mansoni* infection intensity was significantly associated with enlargement. For a doubling of *S. mansoni* egg counts, the relative risk of moderate enlargement increased by 4% and severe enlargement by 8% (per unit increase in log-transformed egg count: moderate enlargement RRR 1.06, 95% CI 1.01–1.12; severe enlargement RRR 1.12, 95% CI 1.04–1.20). Children with malaria had a 2.74-fold increase in the relative risk of moderate enlargement (95% CI 2.08–3.61) and a 3.96-fold increase in relative risk for severe enlargement (95% CI 2.64–5.92) compared to those without malaria. Additionally, the reported use of antimalarials in the past month was associated with a 51% increase in the risk of severe splenic enlargement (RRR 1.51; 95% CI 1.04–2.20).

**Fig 2.**
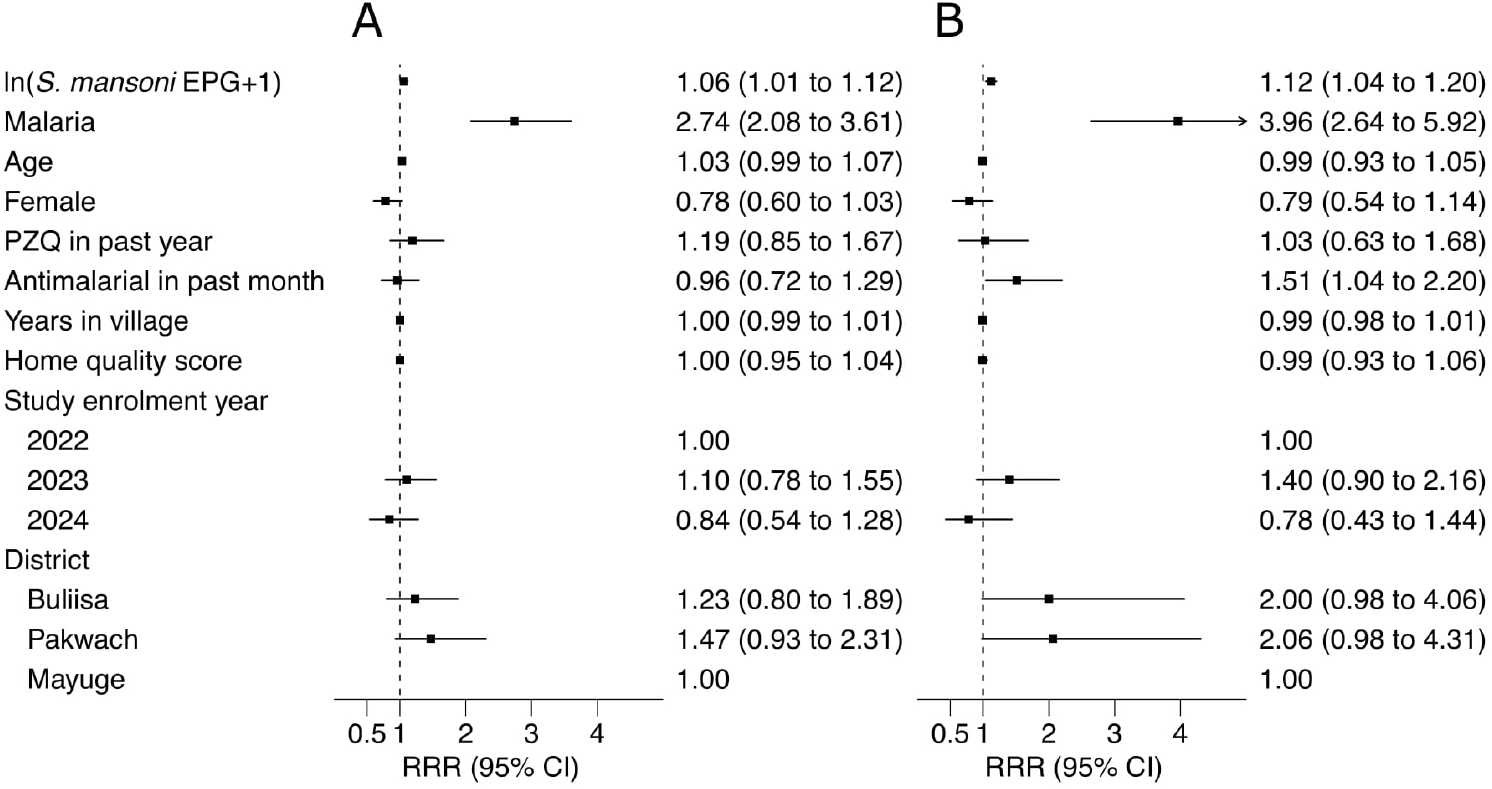
Determinants of spleen dimensions in children. Plots show RRRs with 95% CIs. **A** Moderately enlarged. **B** Severely enlarged. GVIF was < 5.

**Fig 3.**
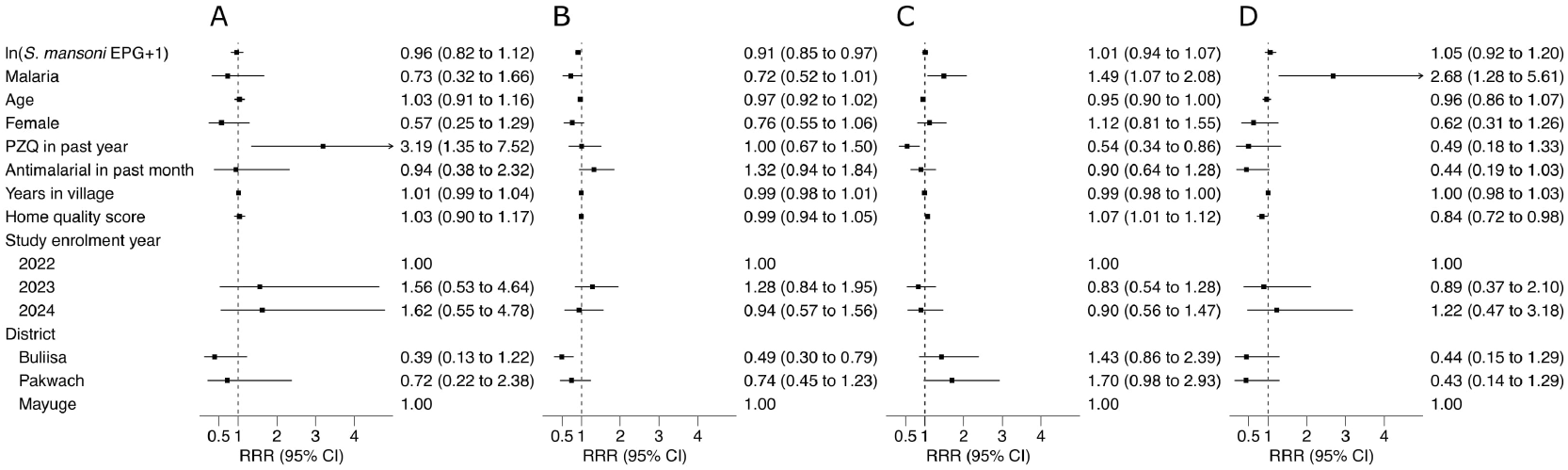
Determinants of left liver lobe abnormality in children. Plots show RRR with 95% CIs. **A** Severely shrunken. **B** Moderately shrunken. **C** Moderately enlarged. **D** Severely enlarged. GVIF was < 5.

**Fig 4.**
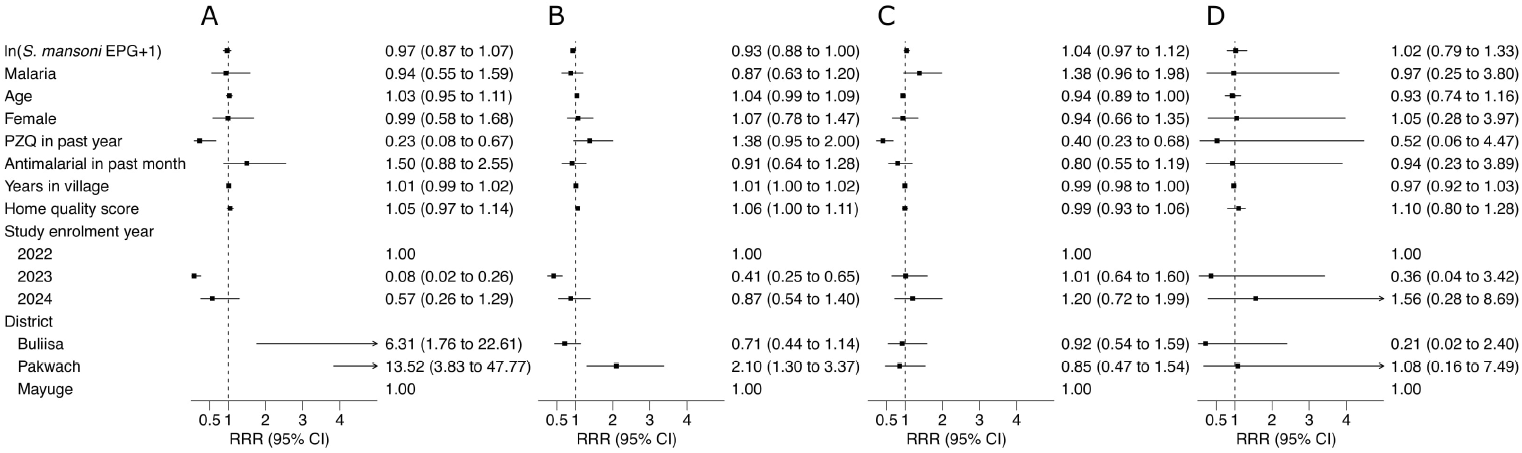
Determinants of right liver lobe dimensions in children. Plots show RRR with 95% CIs. **A** Severely shrunken **B** Moderately shrunken **C** Moderately enlarged **D** Severely enlarged. GVIF was < 5.

Regarding liver organometry in children, associations with *S. mansoni* intensity were unclear. Higher infection intensity was associated with a reduced risk of moderate left liver lobe shrinkage (RRR 0.91; 95% CI 0.85–0.97), but not severe left liver lobe shrinkage (RRR 0.96; 95% CI 0.82–1.12).

Having malaria was associated with a 49% increase in the risk of moderate left lobe enlargement (RRR 1.49; 95% CI 1.07–2.08) and more than a two-fold increase in the risk of severe enlargement (RRR 2.68; 95% CI 1.28–5.61). Praziquantel treatment in the past year through MDA showed inconsistent associations with liver morbidity. Treatment history was identified as a significant correlate of severe left liver lobe shrinkage, associated with a 3.19-fold increase in relative risk (95% CI 1.35–7.52). Conversely, for the right liver lobe, treatment was associated with a 60% reduction in the risk of moderate enlargement (RRR 0.40; 95% CI 0.23–0.68). Living in Pakwach district was associated with a 2.10-fold increase in the relative risk of moderate shrinkage (95% CI 1.30–3.37) and a 13.52-fold increase for severe shrinkage (95% CI 3.83–47.77) as compared to living in Mayuge district. Similarly, residence in Buliisa increased the risk of severe right lobe shrinkage by over six times (RRR 6.31; 95% CI 1.76–22.61) as compared to Mayuge district.

### Determinants of abnormal organometry for adults

In adults, the determinants of organometry are shown in Figures 5, 6, and 7. For splenic morbidity, HIV infection was associated with a 2.71-times increase in the relative risk of moderate enlargement (95% CI 1.20–6.10) and a 3.62-times increase for severe enlargement (95% CI 1.58–8.33). Conversely, antiretroviral therapy (ART) use was associated with a 64% reduction in the relative risk of moderate enlargement (RRR 0.36; 95% CI 0.14–0.94) and a 71% reduction in the relative risk of severe enlargement (RRR 0.29; 95% CI 0.10–0.82). Demographic and geographic factors also contributed to splenic outcomes; occupation as a fisherman was associated with a two-times increase in the risk of severe enlargement (RRR 2.05; 95% CI 1.18–3.54) as compared to other or no occupations, while female sex was associated with lower relative risk of severe enlargement (RRR 0.59; 95% CI 0.36–0.97). Living in Pakwach district was strongly associated with a 5.19-times increase in severe enlargement compared to living in Mayuge (95% CI 2.77–9.74).

**Fig 5.**
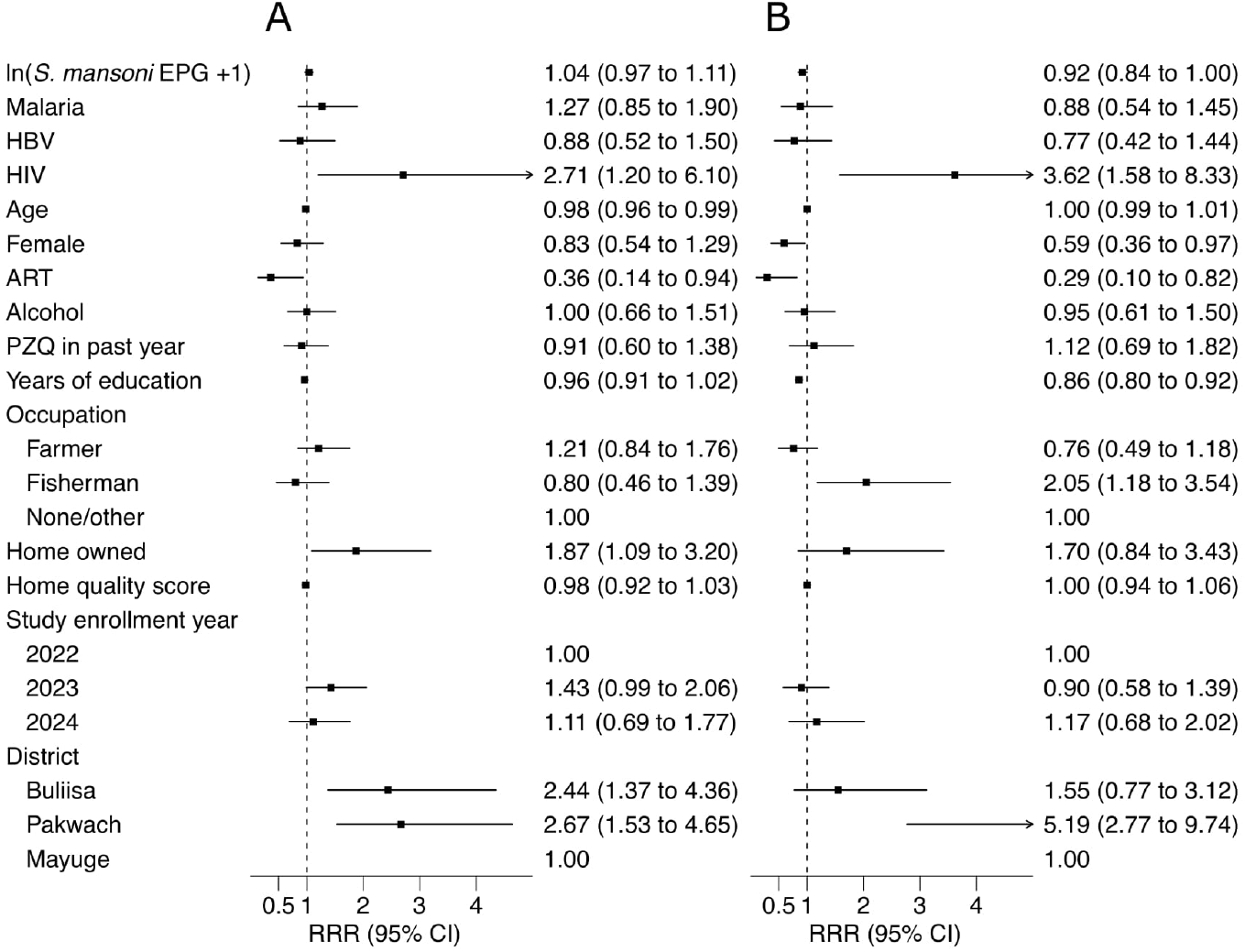
Determinants of spleen dimensions in adults. Plots show RRRs with 95% CIs. **A** Moderately enlarged. **B** Severely enlarged. GVIF remained < 5.

**Fig 6.**
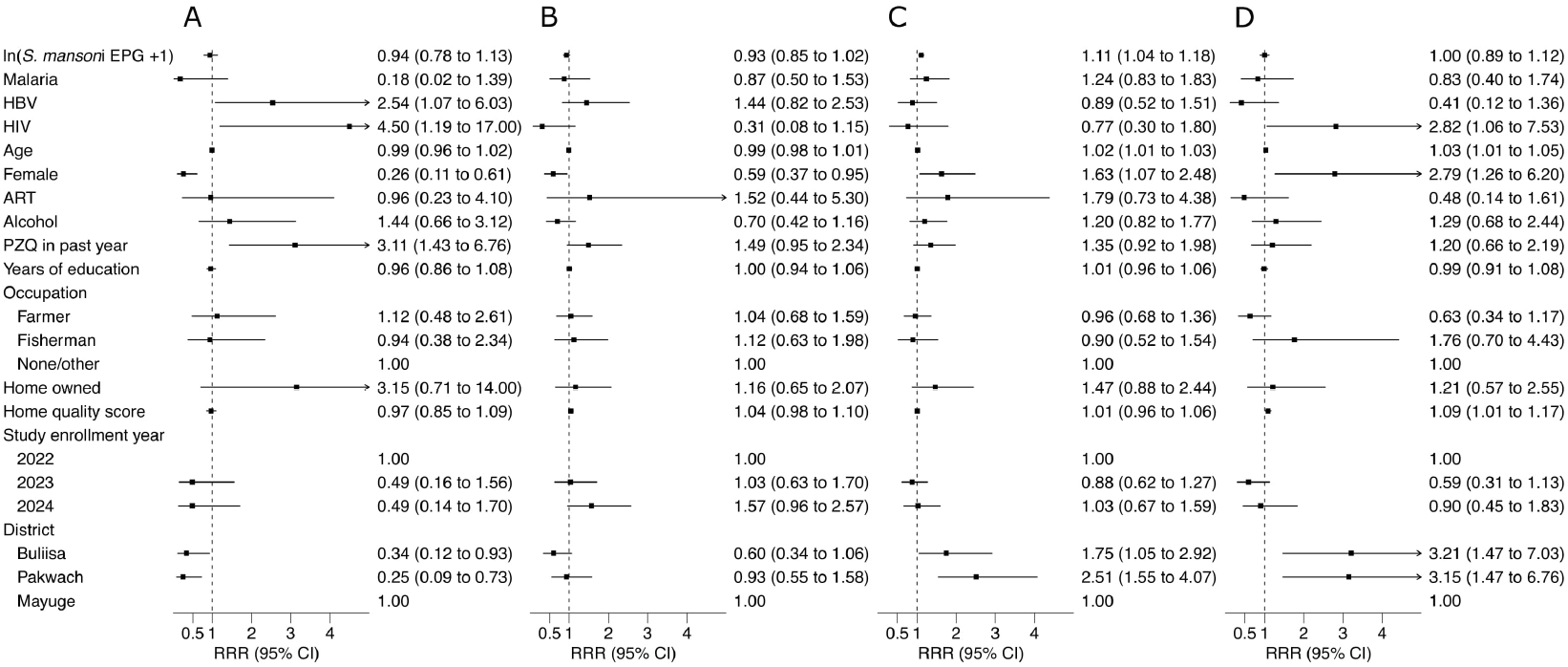
Determinants of left liver lobe dimensions in adults. Plots show RRRs with 95% CIs. **A** Severely shrunken. **B** Moderately shrunken. **C** Moderately enlarged. **D** Severely enlarged. GVIF remained within acceptable limits (< 5).

**Fig 7.**
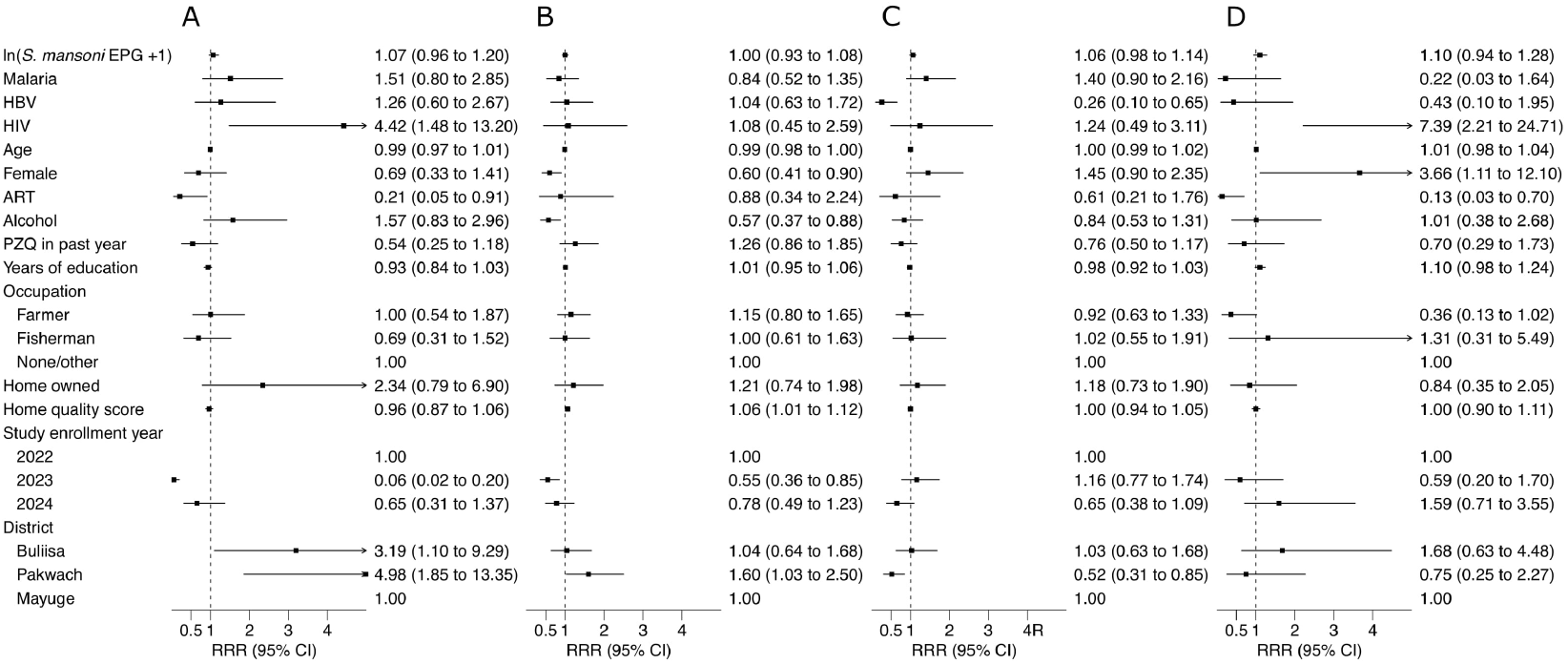
Determinants of right liver lobe dimensions in adults. Plots show RRRs with 95% CIs. **A** Severely shrunken. **B** Moderately shrunken. **C** Moderately enlarged. **D** Severely enlarged. GVIF was < 5.

Regarding the left liver lobe, for a doubling of *S. mansoni* egg counts, the relative risk of moderate enlargement increased by 8% (per unit increase in log-transformed egg count: RRR 1.11; 95% CI 1.04–1.18). Viral infections were significant correlates of severe shrinkage, with HIV infection associated with a 4.50-times increase in relative risk (95% CI 1.19–17.00) and HBV infection with a 2.54-times increase in relative risk (95% CI 1.07–6.03). Praziquantel use in the past year from MDA also was associated with a 3.11-times increase in the relative risk of severe shrinkage (95% CI 1.43–6.76). Female sex was a significant predictor for hepatomegaly, associated with increased relative risks for both moderate (RRR 1.63; 95% CI 1.07–2.48) and severe enlargement (RRR 2.79; 95% CI 1.26–6.20). Additionally, living in Pakwach or Buliisa was strongly associated with both moderate and severe left lobe enlargement as compared to living in Mayuge district.

For the right liver lobe, HIV infection was strongly positively associated with increased relative risk of severe shrinkage (RRR 4.42, 95% CI 1.48–13.20) and severe enlargement (RRR 7.39, 95% CI 2.21–24.71). Similarly, for the left lobe, female sex was associated with a 3.66-times increase in the risk of severe enlargement (95% CI 1.11–12.10). Similar to the left lobe, living in Pakwach corresponded to a nearly five-times increase (RRR 4.98; 95% CI 1.85–13.35) and living in Buliisa to a three-times increase (RRR 3.19; 95% CI 1.10–9.29) in relative risk of severe shrinkage when compared to living in Mayuge district.

### Population attributable fractions (PAFs)

PAFs for significant infection associations with organometry are presented in Table 3. In children, moderate (PAF 30.8%; 95% CI 21.2–41.0) and severe splenic enlargement (PAF 46.7%; 95% CI 31.7–61.3) were often attributable to malaria. Nearly a quarter of cases of splenomegaly also were attributable to *S. mansoni* (severe enlargement PAF 23.6%; 95% CI 5.8–40.7). Over 43% of severe left liver lobe enlargement was attributable to malaria (95% CI 13.9–69.9), while moderate enlargement was less attributable to malaria (PAF 17%, 95% CI: 3.9–29.2) of cases.

**Table 3.**
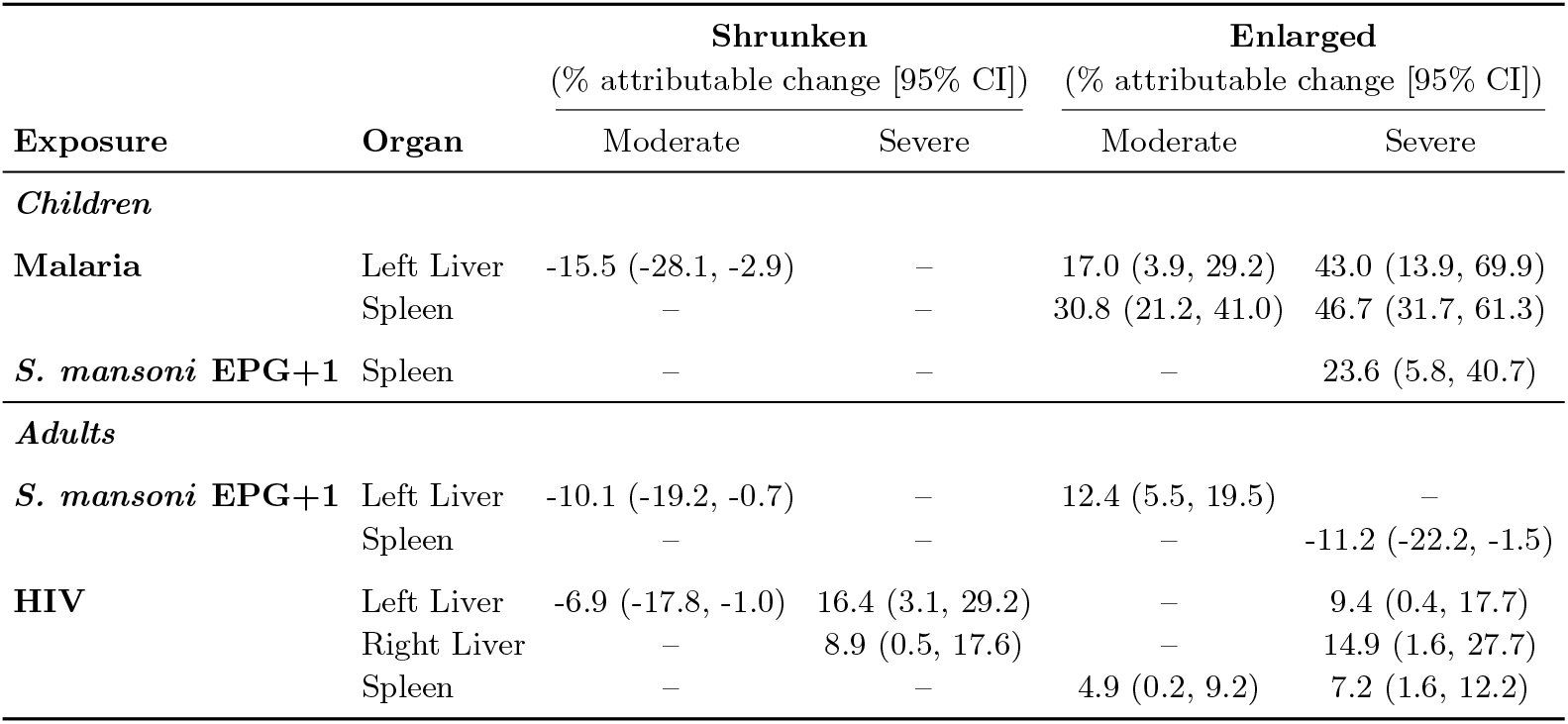
Estimated Population Attributable Fraction in organometry by infection. Values represent the estimated proportion of organ abnormalities statistically attributable to each specific infection using G-computation. All reported values correspond to exposures with significant coefficients (*p* < 0.05) in multivariable models. 95% CIs were derived from bootstrap resampling (n=1000). Positive values indicate the percentage of the outcome prevalence that is statistically attributable to the infection (i.e., the expected reduction in prevalence if the infection were removed). Negative values indicate that the infection is associated with a lower percentage of that specific category.

| Exposure | Organ | Shrunken |  | Enlarged |  |
| --- | --- | --- | --- | --- | --- |
|  |  | (% attributable change [95% CI]) |  | (% attributable change [95% CI]) |  |
|  |  | Moderate | Severe | Moderate | Severe |
| *Children* |  |  |  |  |  |
| Malaria | Left Liver | -15.5 (-28.1, -2.9) | – | 17.0 (3.9, 29.2) | 43.0 (13.9, 69.9) |
|  | Spleen | – | – | 30.8 (21.2, 41.0) | 46.7 (31.7, 61.3) |
| *S. mansoni* EPG+1 | Spleen | – | – | – | 23.6 (5.8, 40.7) |
| *Adults* |  |  |  |  |  |
| *S. mansoni* EPG+1 | Left Liver | -10.1 (-19.2, -0.7) | – | 12.4 (5.5, 19.5) | – |
|  | Spleen | – | – | – | -11.2 (-22.2, -1.5) |
| HIV | Left Liver | -6.9 (-17.8, -1.0) | 16.4 (3.1, 29.2) | – | 9.4 (0.4, 17.7) |
|  | Right Liver | – | 8.9 (0.5, 17.6) | – | 14.9 (1.6, 27.7) |
|  | Spleen | – | – | 4.9 (0.2, 9.2) | 7.2 (1.6, 12.2) |

In adults, severe organometry outcomes were attributable to HIV. 16.4% of severe left liver lobe shrinkage was attributable to HIV (95% CI: 3.1–29.2), with 8.9% (95% CI: 0.5–17.6) of severe right lobe shrinkage and 14.9% (95% CI: 1.6–27.7) of severe right lobe enlargement attributable to HIV. *S. mansoni* accounted for 12.4% of moderate left liver lobe enlargement (95% CI: 5.5–19.5) of cases. Regarding the spleen, 7.2% of severe enlargement was attributable to HIV (95% CI: 1.6–12.2).

### Sensitivity analyses and comparisons to WHO Niamey reference tables

Alternative models utilizing collapsed outcome categories (shrunken, normal, enlarged) yielded results largely consistent with the primary adjusted models (Supplementary Figures S1–S6). A sensitivity analysis substituting malaria status with continuous log-transformed parasite density reconfirmed the dose-dependent relationship between malaria and splenomegaly in children (Supplementary Figure S7). For the outcome of hepatosplenomegaly, defined as any concurrent enlargement of both the liver and spleen, determinants mirrored those of the individual organs (Supplementary Figures S8 and S9).

For both children and adults, the WHO Niamey reference standards systematically underestimated the prevalence of abnormal liver organometry while overestimating the prevalence of splenomegaly (Supplementary Table S7). Using reference standards from the Niamey Protocol identified approximately half the prevalence of severe left liver lobe morbidity compared to SchistoTrack reference standards constructed here and classified only 0.1% of the population as having severe right liver lobe enlargement compared to 1.6% internally. Conversely, regarding the spleen, the Niamey Protocol references produced prevalence estimates for enlargement nearly double those of the internal reference (49.4% vs 26.0%).

## Discussion

Clinical guidance for hepatosplenic schistosomiasis is needed to enable morbidity surveillance and case management. We investigated the relationship between current *S. mansoni* infection intensity and abnormal organometry outcomes within the context of malaria, HIV and HBV co-endemic infection across 3121 children and adults within the SchistoTrack cohort. *S. mansoni* infections were associated with abnormal organometry in both children and adults after controlling for malaria, HIV, and HBV co-endemic infections.

Despite severe splenomegaly in children being largely attributable to malaria (PAF 46.7%), *S. mansoni* infection intensity remained positively associated with moderate splenomegaly in children after controlling for malaria. Furthermore, 23.6% of severe splenomegaly was attributable to *S. mansoni* intensity. The WHO Niamey Protocol [21] recommends excluding the assessment of splenomegaly for schistosomiasis classification in malaria-endemic areas. This guidance is not differentiated for children versus adults. Our study challenges this suggestion as schistosome infections remain associated with splenomegaly in children despite the co-endemicity of malaria. Given the high spatial overlap of schistosome and malaria infections and high prevalence of coinfections [34–36], following the current WHO Niamey protocol guidance [21] would result in gross under-estimations of schistosomiasis-related morbidities. Future revisions of the WHO Niamey Protocol should recommend that splenomegaly be assessed in malaria-endemic areas, while controlling for malaria co-infections in children only.

In children, only malaria was positively correlated to abnormal liver organometry, specifically for left liver lobe enlargement. This finding is notable given that the dominant malaria species in our study area is *Plasmodium falciparum* [31], which is not typically associated with chronic hepatic phase pathology in its life cycle. This contrasts with historical studies [37, 38] suggesting malarial hepatomegaly is common, though often transient. Given the low prevalence of severe liver enlargement in children (2.5%), this result should be interpreted with caution. Future research is needed to understand if hepatomegaly should be considered a clinical marker of chronic *P. falciparum* exposure in children.

Abnormal liver organometry was observed more often in adults than children with inconsistent associations with *S. mansoni* infection and clear associations with HIV and HBV. *S. mansoni* infection only showed a positive association with moderate left lobe enlargement. This association may be an indicator of early morbidity, as it has been suggested in Brazilian hospital-based studies that schistosome fibrosis and pathogenesis are first observed in the left liver lobe [39–41]. This heterogeneity may be attributable to anatomical differences in blood flow from the portal vein, which differentially expose liver lobes to parasite eggs and gut-derived toxins [41].

There were associations of HIV and HBV with abnormal liver organometry. HBV was associated with 2.54 higher RRR of severely shrunken left liver lobes. While HBV is a known cause of cirrhosis, it is unclear why this effect would be isolated to the left lobe, warranting further investigation. HIV was consistently associated with severe outcomes of both liver shrinkage and hepatomegaly in both the left and right liver lobes. Hepatic disorders are common clinical findings among children [42] and adults [43] living with HIV: mono-infection is associated with a higher incidence of non-alcoholic fatty liver disease, non-alcoholic steatohepatitis and hepatic fibrosis [44], and enhances the association of alcohol consumption with related liver pathology [45]. Antiretroviral therapy and HBV treatment can themselves be associated with hepatic pathology. Abdominal tuberculosis and other opportunistic infections may also contribute to abnormalities in people with undiagnosed or advanced HIV infection. Anjorin et al. [3] demonstrated that HIV infection significantly increased the likelihood of schistosomal PPF, independent of current *S. mansoni* infection status. Furthermore, Zhi et al. [2] identified HIV as a key driver of PPF and hepatosplenic multimorbidity using Bayesian multitask machine learning, reinforcing the hypothesis that viral co-infections accelerate the transition from inflammatory morbidity to chronic fibrosis for schistosomiasis-specific fibrosis patterns. While early schistosomal morbidity is believed to manifest in granulomatous-induced inflammation leading to hepatomegaly [46, 47], advanced fibrosis related to PPF due to the condensed structure of the collagen matrix can result in reduced liver volume [46, 48], perhaps even mimicking the atrophic outcome of viral cirrhosis though cirrhosis is assumed to be uncommon for hepatosplenic schistosomiasis [8, 48, 49]. A recent meta-analysis [50] found that co-infections with HBV increased the odds of cirrhosis when compared to singular infections, further supporting that viral-parasitic interactions (here with past schistosome infection) might be driving the liver pathology as opposed to singular infections. Our findings might be consistent with an interaction between HIV and past schistosome infection, however it was not possible to assess this directly in this analysis. Future work is needed to assess HIV and schistosome interactions and ideally prospectively to know the sequence of exposures for untangling interactions from potential immune priming. Our findings underscore a critical gap in current MDA strategies, which typically focus on *S. mansoni* in isolation. HIV and HBV should be considered when developing morbidity assessment tools for schistosomiasis-related morbidity in adults.

We observed an unusual positive association between praziquantel treatment in the past year through MDA and severe liver shrinkage in adults and children. This association likely reflects reverse causality, where participants with established morbidity or symptoms are more likely to seek or be targeted for treatment, rather than a direct pathological effect of the drug. Regardless, praziquantel through MDA did show inconsistent associations and no clear protective pattern. This finding contrasts with pre-MDA studies in children only suggesting praziquantel can reverse early hepatosplenic morbidity [24]. Our results support recent analyses demonstrating persistent morbidity that is not amenable to preventive chemotherapy despite repeated MDA [2, 3, 14]. Studies are currently underway within the SchistoTrack cohort to evaluate morbidity resolution prospectively [26].

Spatial variation exists for the prevalence of abnormal organometry. Western districts (Pakwach and Buliisa) had the highest burden of severe liver shrinkage and splenic enlargement compared to the eastern district of Mayuge. Pakwach district specifically had the highest burden of abnormalities in both children and adults. While Pakwach has historically faced challenges with health infrastructure, recent analyses of health access in this cohort indicate that healthcare-seeking behaviour is not significantly lower in Pakwach compared to other districts [26]. Schistosome infection prevalence as well as prevalence for malaria, HBV, and HIV varied by district, but would have been accounted for in our models and did not negate strong district effects. Future research is needed to determine whether unmeasured exposures distinct to the Albert Nile region or specific genetic factors are driving these disparities.

Methodologically, our study highlights limitations in the fixed height-bin approach currently employed by the Niamey Protocol [21], which relies on external reference values derived from a Senegalese population [33]. The application of these external standards to our cohort resulted in systematic diagnostic error: the Niamey standards classified half the population as having splenomegaly, a gross overestimation likely reflecting the disparity in background malaria endemicity between the populations. Conversely, the reference values from the Senegalese study systematically underestimated severe liver shrinkage (1.2% vs 2.2% identified internally) and severe right lobe enlargement (0.1% vs 1.6%). This discordance is likely driven by the limitations of the original reference dataset, which was generated in a pre-MDA context with a limited age range and exclusion criteria that did not account for HIV or HBV infections. By contrast, our internal references were derived from a larger, contemporary population with rigorous screening for co-endemic viruses and parasites, utilizing height deciles to provide a more granular standardization of organometry. Updated reference tables are needed to assess abnormal organometry. Future research should determine whether a global reference table, similar to WHO child growth standards, can be developed or if country-specific tables are necessary. The reference table generated in our study should be tested in other contexts; if generalizable, it could be used where local data is insufficient.

The strengths of our study lie in its comprehensive assessment of risk factors, large sample and age range, robust models, construction of PAFs, and detailed reference tables. Study limitations concern the focus on a single timepoint and focus on clinical interpretability rather than predictive modelling. The cross-sectional design does not allow for causal understanding of pathogenesis, particularly regarding the temporal sequence of infection exposures and changes in organometry over time. Prospective analyses are needed to assess the predictive performance of abnormal organometry for more severe schistosomiasis-related outcomes such as portal hypertension or variceal bleeding.

## Conclusion

Accurately defining routine clinical indicators of hepatosplenic schistosomiasis is necessary to enable the development of morbidity diagnosis and clinical management guidelines in schistosomiasis-endemic areas. *S. mansoni* remains correlated with splenomegaly despite co-endemic malaria, but as HIV appears associated with more severe hepatic organometry, future research must investigate potential interactions between these pathogens. Organometry should be considered in routine health system indicators suggested for schistosomiasis-morbidity monitoring and, if the reference table provided here is generalizable to other contexts, our study can be used to inform assessments of abnormal organometry in large-scale surveillance surveys to improve the estimation of schistosomiasis-related disease burden in SSA.

## Declarations

### Ethical Approval

Ethical approval was granted by the Oxford Tropical Research Ethics Committee (OxTREC 509-21), the Vector Control Division Research Ethics Committee of the Uganda Ministry of Health (VCDREC146), and the Uganda National Council of Science and Technology (UNCST HS 1664ES). Written informed consent was obtained for all adult participants. For children, written informed consent was provided by a parent or guardian, alongside informed assent from the child.

### Availability of data and materials

The raw data are protected and are not available due to data privacy and ethics restrictions. All relevant metadata and supplementary material are provided within the manuscript. The code to reproduce the pipeline is available as supplementary material.

### Conflicts of interest

The authors declare no conflicts of interest.

### Funding

A Nuffield Department of Population Health Post-MSc award was received by KS. A DPhil scholarship was awarded from the Nuffield Department of Population Health to MML. Grants from the Wellcome Trust Institutional Strategic Support Fund (204826/Z/16/Z), NDPH Pump Priming Fund, Robertson Foundation, and UKRI EPSRC Award (EP/X021793/1) were awarded to GFC. For the purpose of Open Access, the author has applied a CC-BY public copyright licence to any Author Accepted Manuscript version arising from this submission.

### Author Contributions

Conceptualisation: GFC

Data curation: KS, MML, BNa, VA, BNt, TM, SM, NBK, and GFC.

Formal analysis: KS and MML.

Funding acquisition: GFC.

Investigation: KS, MML, AS, and GFC.

Methodology: KS, MML, GFC.

Project administration: NBK and GFC.

Resources: GFC.

Software: GFC.

Supervision: AS and GFC.

Validation: KS and MML.

Visualisation: KS and MML.

Writing–original draft: KS, MML, and GFC.

Writing–review & editing: KS, MML, BNa, VA, BNt, TM, SM, NBK, AS, and GFC.

## Acknowledgements

We are thankful for the initial input on the scripts to generate the organometry reference tables from Chris Ho. We are grateful for the engagement and participation of all study participants and local communities involved in this research. We extend our thanks to all field teams, auxiliary workers, and community members who contributed to the data collection efforts. We thank the SchistoTrack group for valuable feedback and insights during group meetings and conversations.

## List of abbreviations

ART: Antiretroviral Therapy
CI: Confidence Interval
EPG: Eggs Per Gram
HBV: Hepatitis B Virus
HIV: Human Immunodeficiency Virus
IQR: Interquartile Range
LRT: Likelihood Ratio Test
MDA: Mass Drug Administration
OR: Odds Ratio
OxTREC: Oxford Tropical Research Ethics Committee
PAF: Population Attributable Fraction
PPF: Periportal Fibrosis
RDT: Rapid Diagnostic Test
RRR: Relative Risk Ratio
SD: Standard Deviation
SSA: Sub-Saharan Africa
UNCST: Uganda National Council of Science and Technology
VCDREC: Vector Control Division Research Ethics Committee
GVIF: Generalized Variance Inflation Factor
WASH: Water, Sanitation, and Hygiene
WHO: World Health Organization

## Notes

### Competing Interest Statement

The authors have declared no competing interest.

