## Supplementary File for "*Schistosoma mansoni* infections are associated with hepatosplenic organometry within the context of repeated praziquantel treatment and co-endemic infections of malaria, hepatitis B, and HIV: a population-based study in rural Uganda"

### Supporting information

#### Supplementary Figures

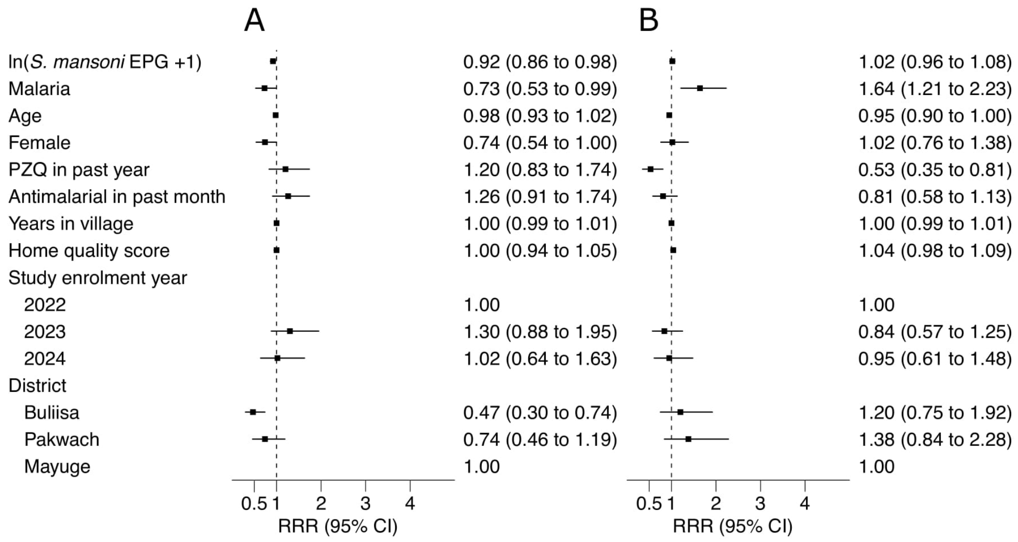


**Fig S1. Determinants of collapsed left liver lobe dimensions in children.** Plots show RRRs with 95% Confidence Intervals. **A** Shrunken. **B** Enlarged.

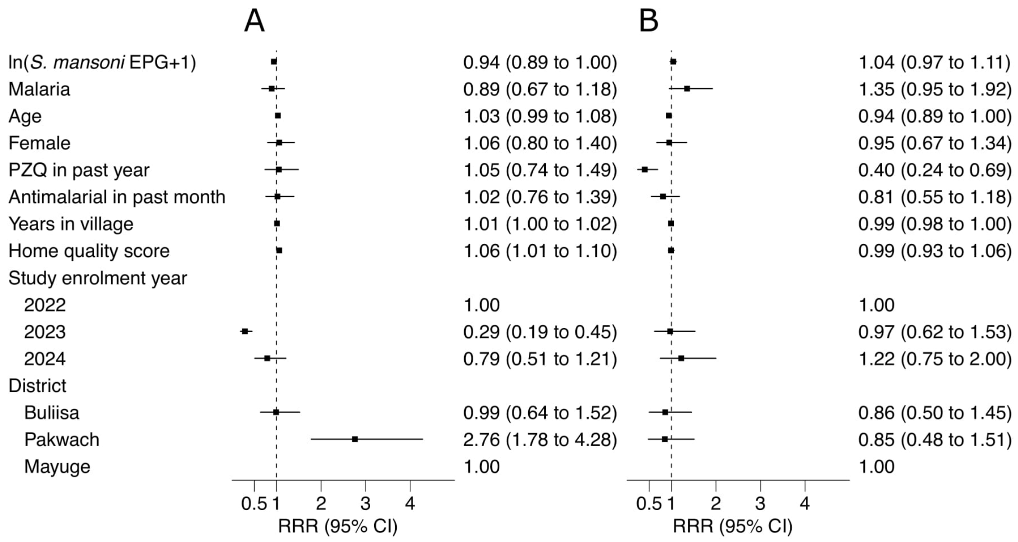


**Fig S2. Determinants of collapsed right liver lobe dimensions in children.** Plots show RRRs with 95% Confidence Intervals. **A** Shrunken. **B** Enlarged.

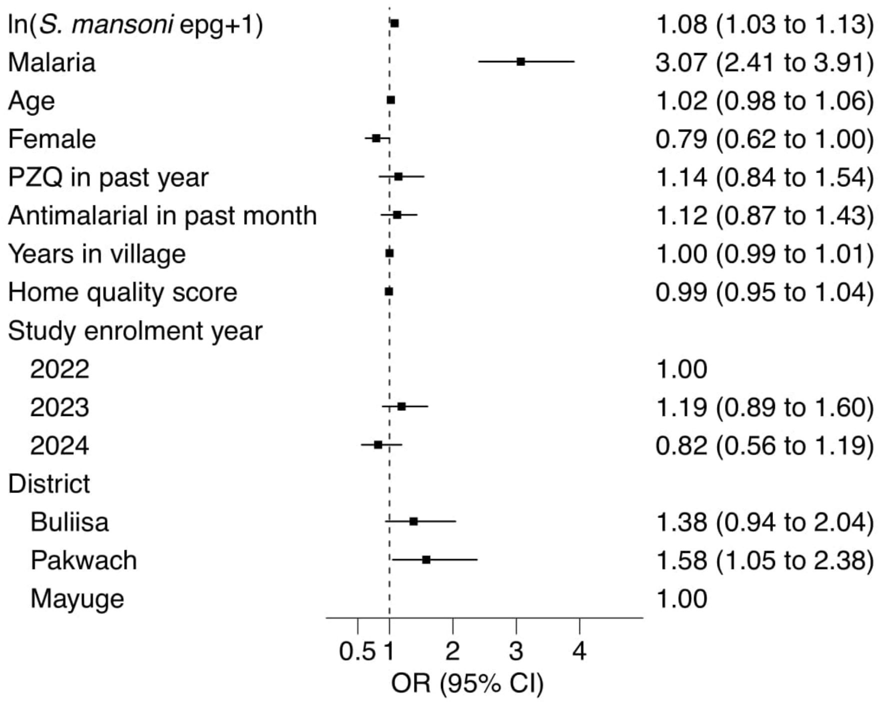


**Fig S3. Determinants of collapsed spleen dimensions in children.** Plots show ORs with 95% Confidence Intervals.

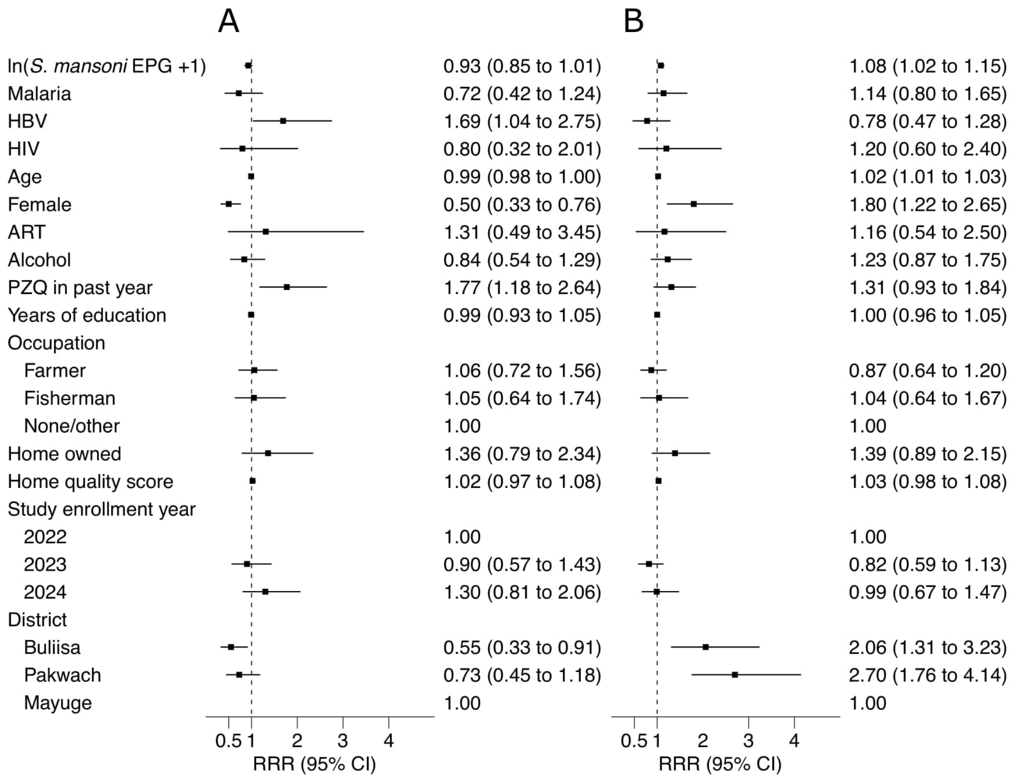


**Fig S4. Determinants of collapsed left liver lobe dimensions in adults.** Plots show RRRs with 95% Confidence Intervals. **A** Shrunken. **B** Enlarged.

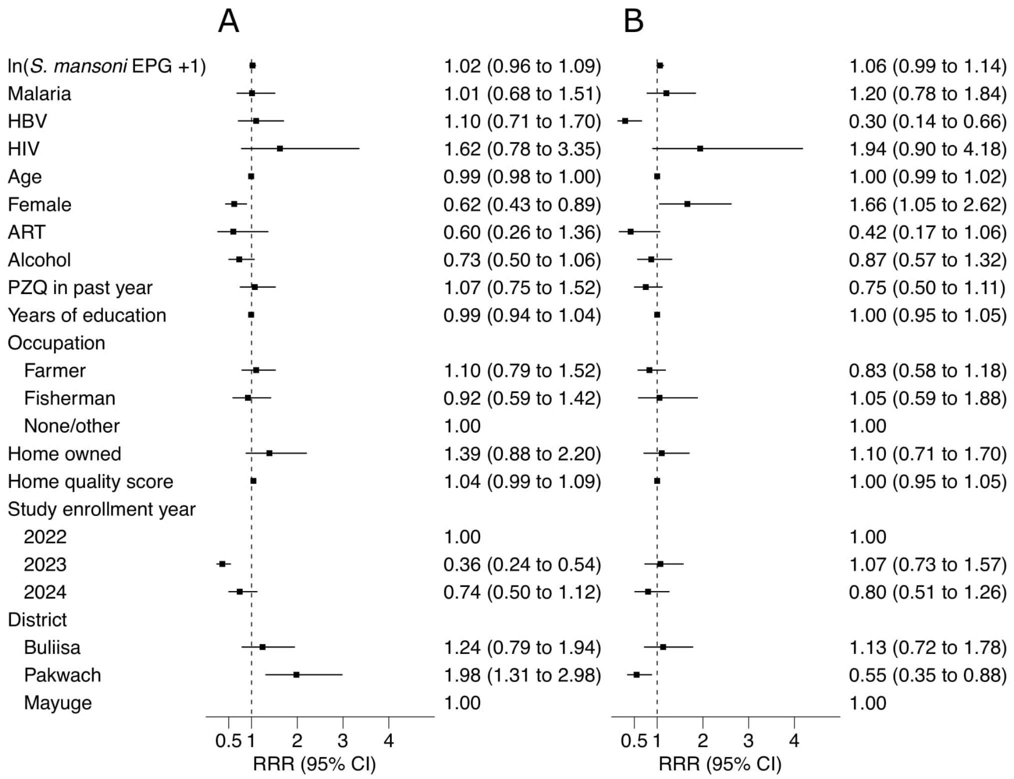


**Fig S5. Determinants of collapsed right liver lobe dimensions in adults.** Plots show RRRs with 95% Confidence Intervals. **A** Shrunken. **B** Enlarged.

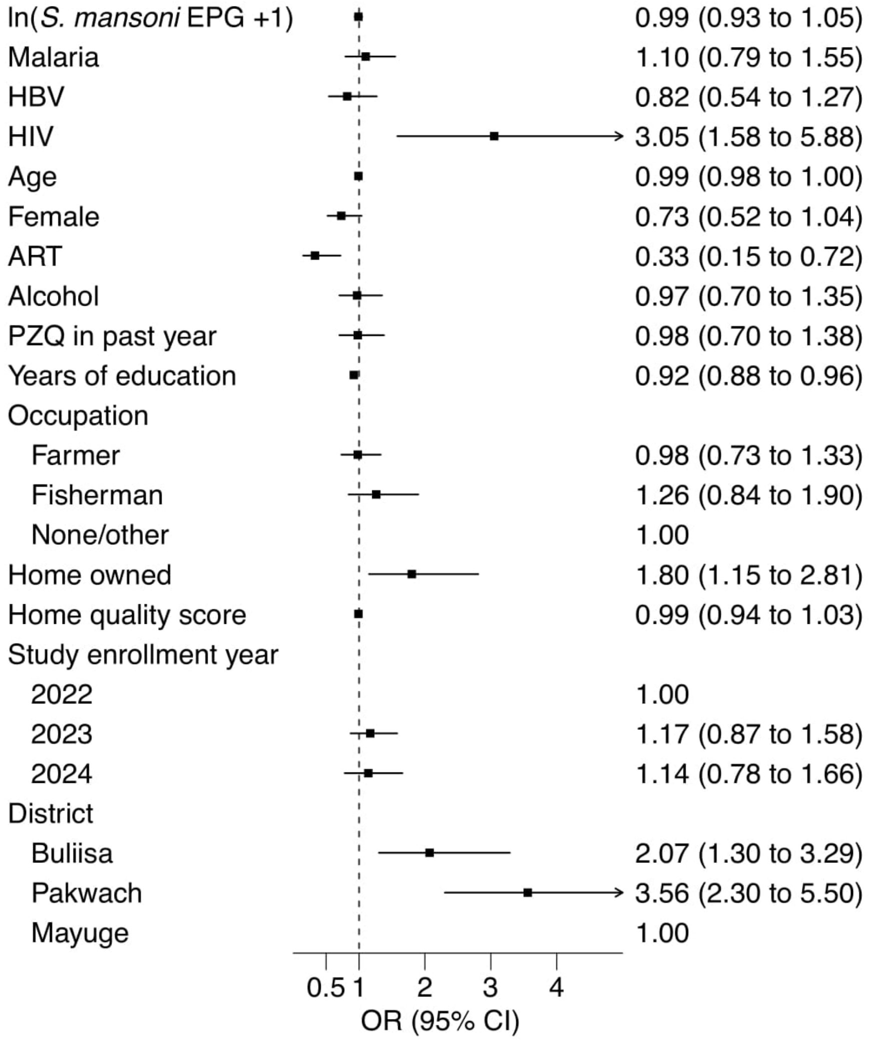


**Fig S6. Determinants of collapsed spleen dimensions in adults.** Plots show ORs with 95% Confidence Intervals.

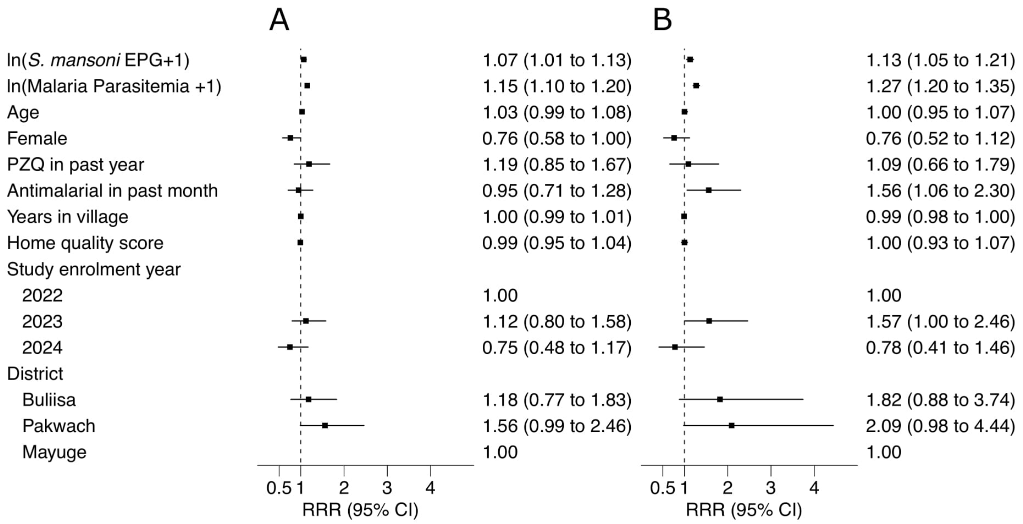


**Fig S7. Determinants of spleen dimensions in children using continuous malaria parasite density.** The model substitutes binary malaria RDT status with natural log-transformed parasite density ($\ln(\text{parasites}/\mu\text{L} + 1)$). Plots show RRRs with 95% Confidence Intervals. **A** Moderately enlarged. **B** Severely enlarged.

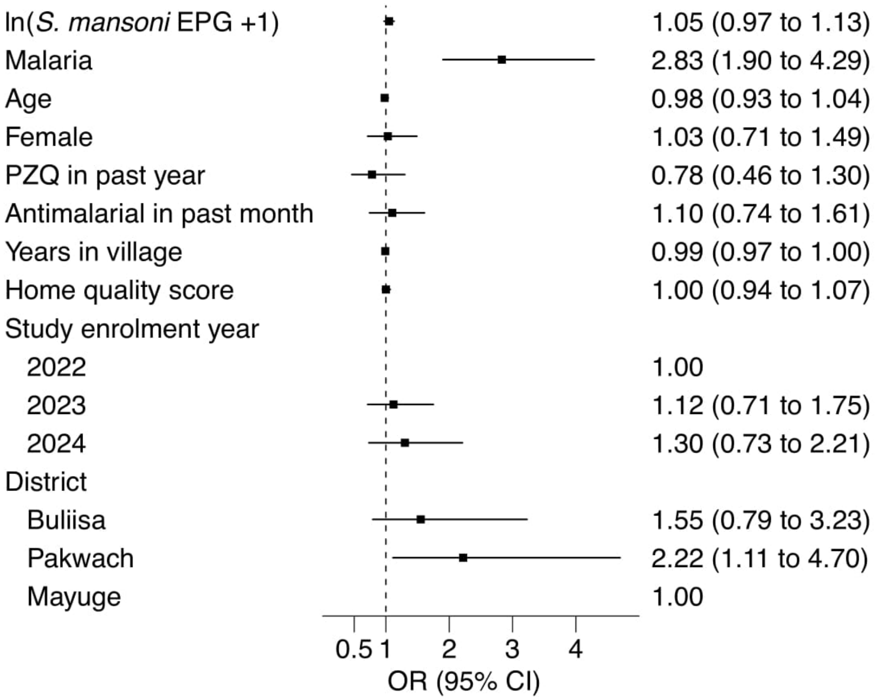


**Fig S8. Determinants of hepatosplenomegaly in children.** Plots show OR with 95% Confidence Intervals for the composite outcome of concurrent liver and splenic enlargement.

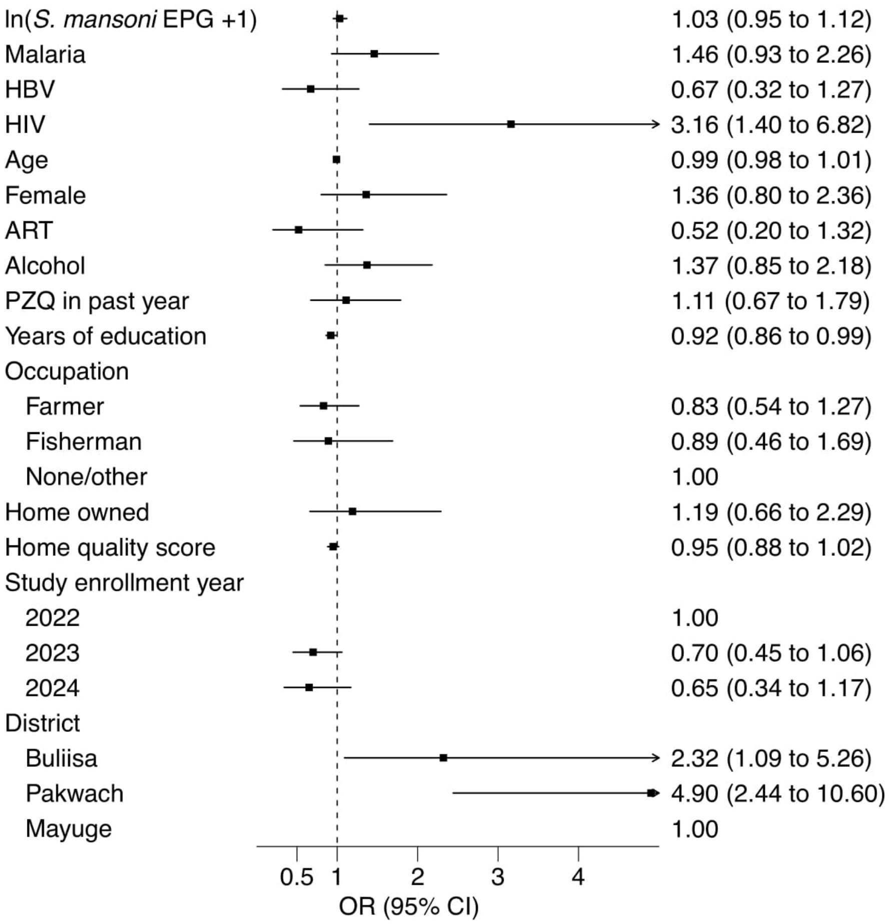


**Fig S9. Determinants of hepatosplenomegaly in adults.** Plots show OR with 95% Confidence Intervals for the composite outcome of concurrent liver and splenic enlargement.

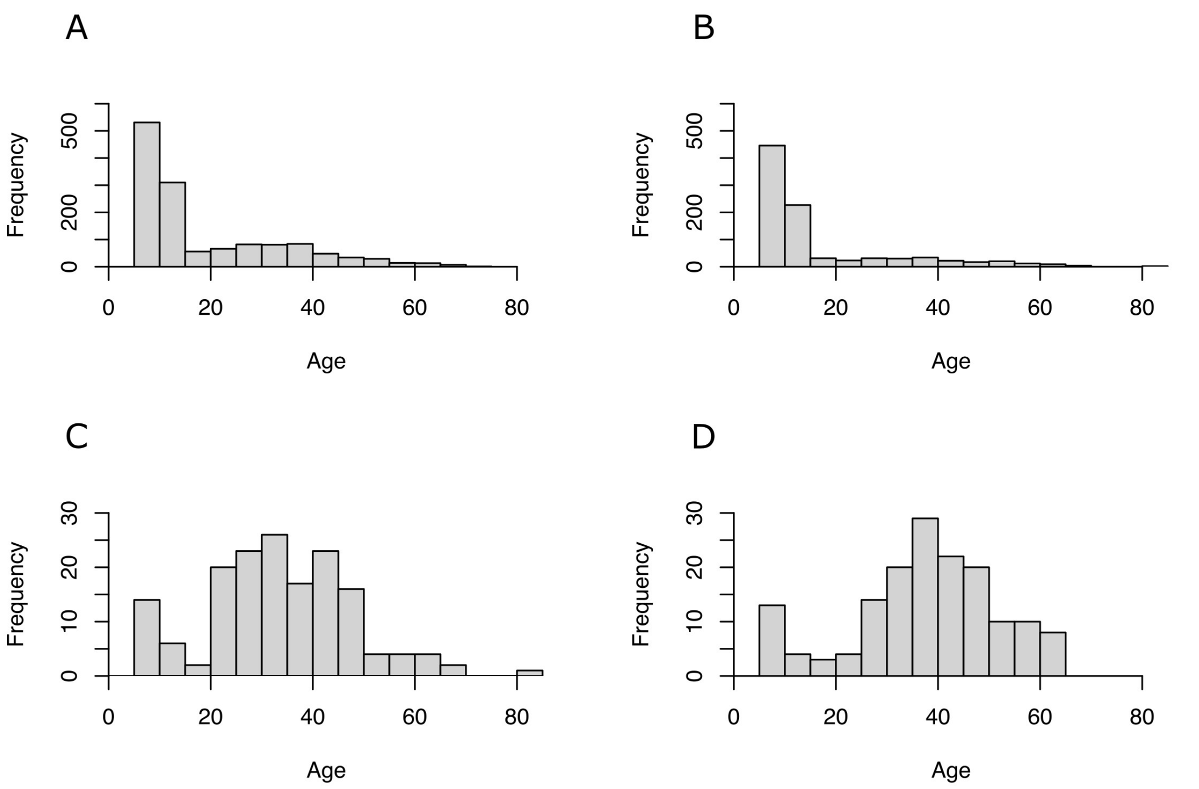


**Fig S10. Frequency of infections by age.** Histograms display the age distribution of positive cases for each infection. **A** *S. mansoni*. **B** Malaria. **C** HBV. **D** HIV.

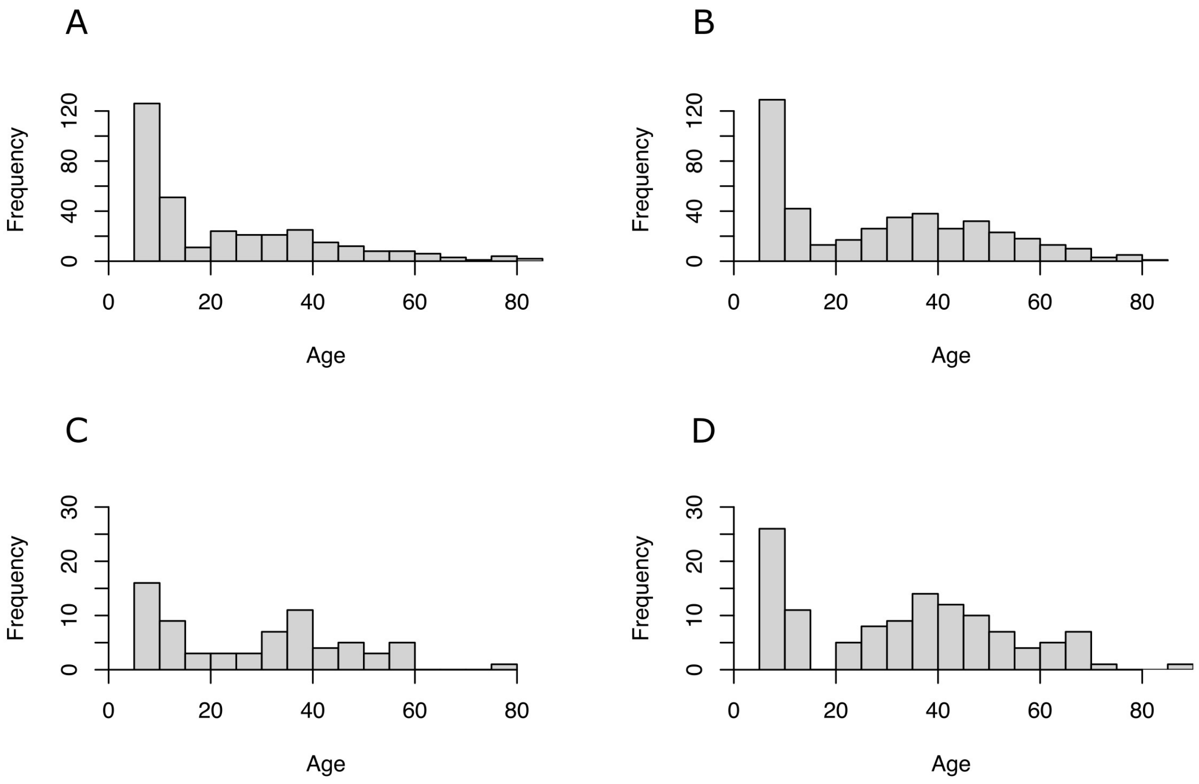


**Fig S11. Frequency of left liver lobe outcomes by age.** **A** Moderately shrunken. **B** Moderately enlarged. **C** Severely shrunken. **D** Severely enlarged.

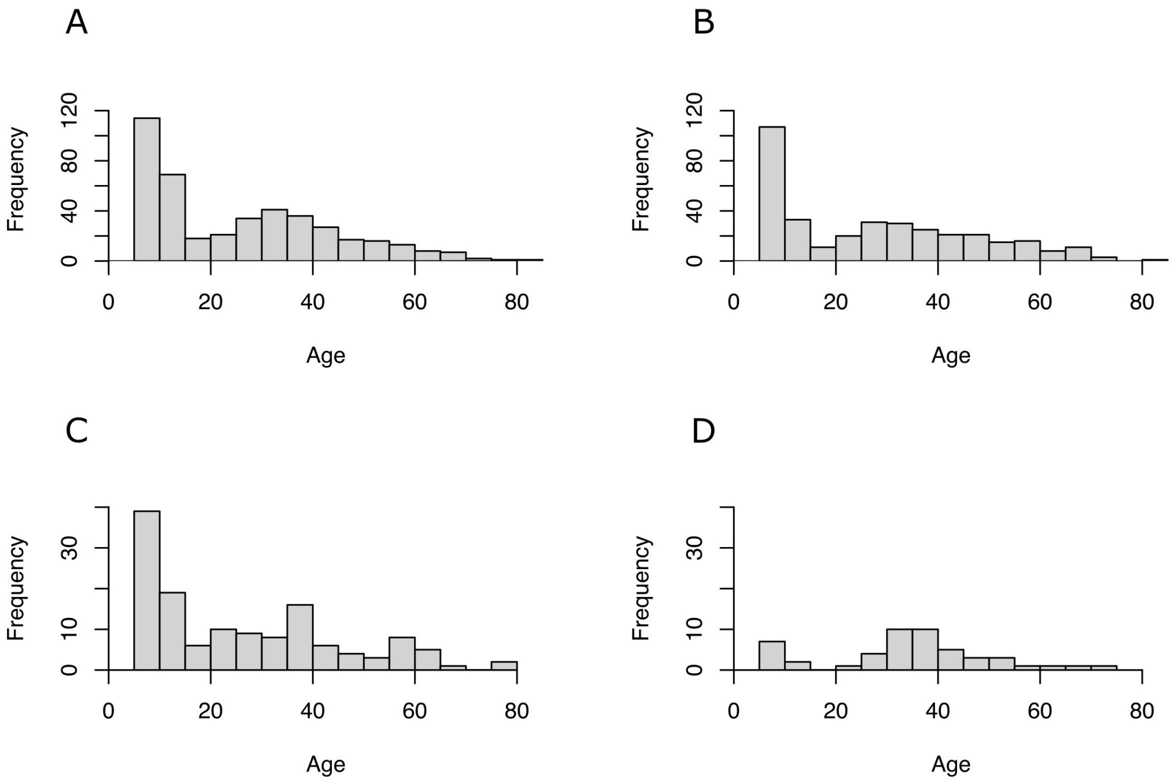


**Fig S12. Frequency of right liver lobe outcomes by age. A** Moderately shrunken. **B** Moderately enlarged. **C** Severely shrunken. **D** Severely enlarged.

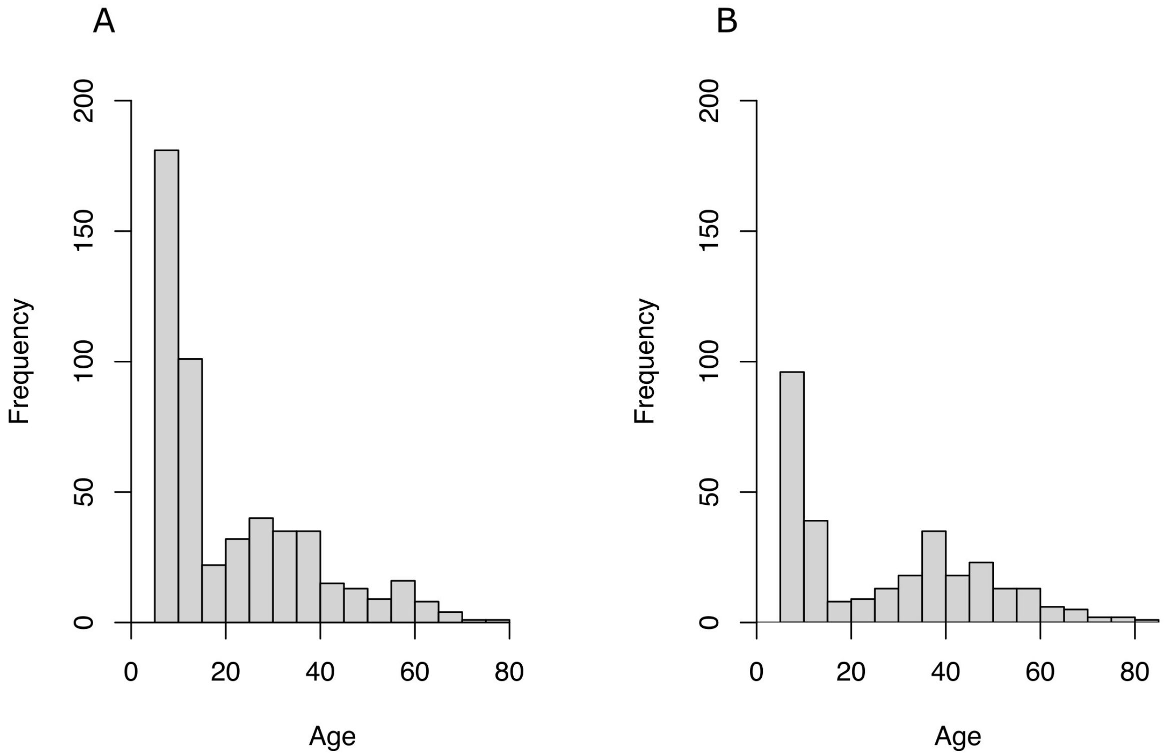


**Fig S13. Frequency of spleen size outcomes by age. A** Moderately enlarged. **B** Severely enlarged.

#### Supplementary Tables

**Table S1. Cross-tabulation of left and right liver lobe classifications.** Data represent the pediatric population (N=1507). $\chi^2 = 203.9$, $df = 16$, $p < 0.001$.

| | Right liver lobe | | | | | |
|---|---|---|---|---|---|---|
| **Left liver lobe** | **SS (-2)** | **MS (-1)** | **Normal (0)** | **ME (1)** | **SE (2)** | **Total** |
| **Severely Shrunken (-2)** | 6<br>(22.2%) | 14<br>(51.9%) | 7<br>(25.9%) | 0<br>(0.0%) | 0<br>(0.0%) | **27**<br>(100%) |
| **Moderately Shrunken (-1)** | 9<br>(4.9%) | 45<br>(24.7%) | 126<br>(69.2%) | 1<br>(0.5%) | 1<br>(0.5%) | **182**<br>(100%) |
| **Normal (0)** | 36<br>(3.3%) | 121<br>(11.2%) | 834<br>(76.9%) | 91<br>(8.4%) | 2<br>(0.2%) | **1084**<br>(100%) |
| **Moderately Enlarged (1)** | 10<br>(5.6%) | 12<br>(6.8%) | 113<br>(63.8%) | 38<br>(21.5%) | 4<br>(2.3%) | **177**<br>(100%) |
| **Severely Enlarged (2)** | 2<br>(5.4%) | 3<br>(8.1%) | 16<br>(43.2%) | 14<br>(37.8%) | 2<br>(5.4%) | **37**<br>(100%) |
| **Total** | **63**<br>(4.2%) | **195**<br>(12.9%) | **1096**<br>(72.7%) | **144**<br>(9.6%) | **9**<br>(0.6%) | **1507**<br>(100%) |

**Table S2. Cross-tabulation of left and right liver lobe classifications (Adults).** Data represent the adult population (N=1614). Values are presented as n (row %). $\chi^2 = 193.58$, $df = 16$, $p < 0.001$.

| | Right liver lobe | | | | | |
|---|---|---|---|---|---|---|
| **Left liver lobe** | **SS (-2)** | **MS (-1)** | **Normal (0)** | **ME (1)** | **SE (2)** | **Total** |
| **Severely Shrunken (-2)** | 8<br>(18.6%) | 19<br>(44.2%) | 15<br>(34.9%) | 1<br>(2.3%) | 0<br>(0.0%) | **43**<br>(100%) |
| **Moderately Shrunken (-1)** | 12<br>(7.7%) | 46<br>(29.5%) | 88<br>(56.4%) | 8<br>(5.1%) | 2<br>(1.3%) | **156**<br>(100%) |
| **Normal (0)** | 38<br>(3.5%) | 131<br>(12.2%) | 767<br>(71.2%) | 126<br>(11.7%) | 16<br>(1.5%) | **1078**<br>(100%) |
| **Moderately Enlarged (1)** | 10<br>(3.9%) | 26<br>(10.2%) | 152<br>(59.8%) | 56<br>(22.0%) | 10<br>(3.9%) | **254**<br>(100%) |
| **Severely Enlarged (2)** | 5<br>(6.0%) | 8<br>(9.6%) | 40<br>(48.2%) | 18<br>(21.7%) | 12<br>(14.5%) | **83**<br>(100%) |
| **Total** | **73**<br>(4.5%) | **230**<br>(14.3%) | **1062**<br>(65.8%) | **209**<br>(12.9%) | **40**<br>(2.5%) | **1614**<br>(100%) |

**Table S3. Lack of selection bias due to missing data.** Baseline characteristics of included vs. excluded participants. P-values calculated via Pearson's Chi-squared test for categorical variables or Wilcoxon rank sum test for continuous variables.

| **Variable** | **Excluded** (N = 181) | **Included** (N = 3121) | **p-value** |
|---|---|---|---|
| *S. mansoni* | 77 (47%) | 1356 (43%) | 0.3 |
| Malaria | 40 (24%) | 908 (29%) | 0.14 |
| HBV | 11 (6.5%) | 162 (5.2%) | 0.4 |
| HIV | 3 (2.5%) | 157 (5.0%) | 0.2 |
| Age | 16 (10, 35) | 21 (9, 40) | 0.3 |
| Sex (female) | 96 (53%) | 1721 (55%) | 0.6 |
| ART | 2 (1.2%) | 123 (4.0%) | 0.075 |
| Praziquantel in past year | 20 (15%) | 683 (22%) | 0.066 |
| Antimalarials in past month | 46 (35%) | 865 (28%) | 0.074 |
| Alcohol | 16 (12%) | 331 (14%) | 0.4 |
| Smoking | 9 (6.6%) | 230 (10.0%) | 0.2 |
| Years of education | 3 (1, 5) | 3 (1, 5) | 0.6 |
| Occupation | | | 0.4 |
| Farmer | 24 (13%) | 528 (17%) | |
| Fisherman | 15 (8.3%) | 221 (7.1%) | |
| None/Other | 142 (78%) | 2372 (76%) | |
| Water contact | 154 (85%) | 2694 (86%) | 0.6 |
| Tribe | | | 0.8 |
| Alur | 118 (65%) | 1936 (62%) | |
| Bagungu | 16 (8.8%) | 297 (9.5%) | |
| Musoga | 17 (9.4%) | 361 (12%) | |
| Other | 30 (17%) | 527 (17%) | |
| Majority religion | 138 (76%) | 2366 (76%) | 0.9 |
| Enrolment year | | | 0.3 |
| 2022 | 106 (59%) | 1978 (63%) | |
| 2023 | 43 (24%) | 713 (23%) | |
| 2024 | 32 (18%) | 430 (14%) | |
| Home quality | 3 (3, 9) | 3 (3, 9) | 0.8 |
| Water purification | 43 (24%) | 700 (22%) | 0.7 |
| Hygiene | 19 (10%) | 262 (8.4%) | 0.3 |
| Sanitation | 103 (57%) | 790 (57%) | > 0.9 |
| Household status | 70 (39%) | 1123 (36%) | 0.5 |
| Years in village | 20 (9, 33) | 18 (8, 30) | 0.3 |
| Home ownership | 144 (80%) | 2742 (88%) | 0.001 |
| District | | | 0.5 |
| Buliisa | 64 (35%) | 1032 (33%) | |
| Mayuge | 36 (20%) | 740 (24%) | |
| Pakwach | 81 (45%) | 1349 (43%) | |

**Table S4. Prevalence of co-infections.** Counts and percentages of participants with specific co-infection pairs stratified by age group.

| **Co-infection** | **Children (n = 1507)** | **Adults (n = 1614)** |
|---|---|---|
| *S. mansoni* - malaria | 437 (29 %) | 74 (5%) |
| *S. mansoni* - HBV | 17 (1%) | 57 (4%) |
| *S. mansoni* - HIV | 14 (1%) | 37 (2%) |
| Malaria - HBV | 8 (1%) | 18 (1%) |
| Malaria - HIV | 6 (<1%) | 18 (1%) |
| HBV - HIV | 0 (0%) | 13 (1%) |

**Table S5. *S. mansoni* status counts by district and age group.** Comparison of negative and positive infection counts across Mayuge, Buliisa, and Pakwach districts.

| | Children | | Adults | |
|---|---|---|---|---|
| District | *S. mansoni* Negative | *S. mansoni* Positive | *S. mansoni* Negative | *S. mansoni* Positive |
| Mayuge | 229 | 106 | 347 | 58 |
| Buliisa | 190 | 317 | 359 | 166 |
| Pakwach | 210 | 455 | 430 | 254 |

**Table S6. Covariate definitions.** Detailed description of variable types and definitions for individual, household, and district level covariates. Abbreviations: ART (antiretroviral therapy), Praziquantel.

| **Variable Name** | **Type** | **Description** |
|---|---|---|
| | ***Individual level*** | |
| **Age** | Continuous | Participant age to nearest year |
| **Sex** | Binary | Participant sex<br>*Male = 0, Female = 1* |
| **Alcohol** | Binary | Self-reported alcohol use (ever or currently)<br>*No = 0, Yes = 1* |
| **Smoking** | Binary | Self-reported smoking (ever or currently)<br>*No = 0, Yes = 1* |
| **ART** | Binary | Self-reported ART use (ever or currently)<br>*No = 0, Yes = 1* |
| **Praziquantel in past year** | Binary | Self-reported treatment with praziquantel in past year<br>*No = 0, Yes = 1* |
| **Antimalarial in past month** | Binary | Self-reported antimalarial treatment in past month<br>*No = 0, Yes = 1* |
| **Education** | Continuous | Self-reported years of education attained |

Continued on next page

**Table S6 – continued from previous page**

| Variable Name | Type | Description |
|---|---|---|
| **Occupation** | Categorical | Self-reported occupation<br>*Fisherman, Farmer, None/Other (reference)* |
| **Water contact** | Binary | Self-reported participation in any activity involving lake water contact<br>*No = 0, Yes = 1* |
| **Tribe** | Categorical | Self-reported participant tribe<br>*Alur, Musoga, Bagungu, Other (reference)* |
| **Majority religion** | Binary | Self-identified religion same as most common religion in village<br>*No = 0, Yes = 1* |
| | **Household level** | |
| **Enrollment year** | Continuous | Year household enrolled in study |
| **Home quality** | Continuous | Sum of scores based on roof, floor, and wall material<br>*Floor: mud (+1), plastic (+2), wood (+3), bricks/cement (+4)*<br>*Wall: mud/sticks (+1), plastic (+2), metal (+3), bricks/cement (+4)*<br>*Roof: grass/papyrus (+1), sticks (+2), plastic (+3), metal (+4)* |
| **Water purification** | Binary | Household uses any technique to make water safer to drink<br>*No = 0, Yes = 1* |
| **Hygiene** | Binary | Household has handwashing facility with soap and water<br>*No = 0, Yes = 1* |
| **Sanitation** | Binary | Household has improved sanitation (private pit/flush latrine)<br>*No = 0, Yes = 1* |
| **Household status** | Binary | Any member of household on local council, religious/tribe leader, village health team, or beach management team<br>*No = 0, Yes = 1* |
| **Years in village** | Continuous | Number of years household head lived in village |
| **Own home** | Binary | Household head owns home<br>*No = 0, Yes = 1* |
| | **District level** | |
| **District** | Categorical | District household located in<br>*Buliisa, Pakwach, Mayuge (reference)* |

**Table S7. Comparison of organometric prevalence using Internal (SchistoTrack) vs. WHO/Niamey protocol reference standards [21].**

| | Internal | | WHO (Niamey) | |
|---|---|---|---|---|
| **Organ / Classification** | **n** | **%** | **n** | **%** |
| **Right liver lobe** | | | | |
| Severely Shrunken | 136 | 4.4 | 97 | 3.1 |
| Moderately Shrunken | 425 | 13.6 | 520 | 16.7 |
| Normal | 2158 | 69.1 | 2388 | 76.5 |
| Moderately Enlarged | 353 | 11.3 | 112 | 3.6 |
| Severely Enlarged | 49 | 1.6 | 4 | 0.1 |
| **Left liver lobe** | | | | |
| Severely Shrunken | 70 | 2.2 | 36 | 1.2 |
| Moderately Shrunken | 338 | 10.8 | 318 | 10.2 |
| Normal (0) | 2162 | 69.3 | 2421 | 77.6 |
| Moderately Enlarged | 431 | 13.8 | 289 | 9.3 |
| Severely Enlarged | 120 | 3.8 | 57 | 1.8 |
| **Spleen** | | | | |
| Severely Shrunken | – | – | 9 | 0.3 |
| Moderately Shrunken | – | – | 26 | 0.8 |
| Normal | 2307 | 73.9 | 1542 | 49.4 |
| Moderately Enlarged | 513 | 16.4 | 903 | 28.9 |
| Severely Enlarged | 301 | 9.6 | 641 | 20.5 |

**Table S8.** Prevalence of Infections by District

| District | Total N | *S. mansoni* | Malaria | HBV | HIV |
|---|---|---|---|---|---|
| Buliisa | 1032 | 483 (46.8%) | 268 (26.0%) | 61 (5.9%) | 54 (5.2%) |
| Mayuge | 740 | 164 (22.2%) | 130 (17.6%) | 21 (2.8%) | 31 (4.2%) |
| Pakwach | 1349 | 709 (52.6%) | 510 (37.8%) | 80 (5.9%) | 72 (5.3%) |
